# Gut microbiota predict development of post-discharge diabetes mellitus in acute pancreatitis

**DOI:** 10.1101/2025.08.12.25333486

**Authors:** Christoph Ammer-Herrmenau, Richard Meier, Kai L Antweiler, Thomas Asendorf, Silke Cameron, Gabriele Capurso, Marko Damm, Linh Dang, Fabian Frost, Jacob Hamm, Albrecht Hoffmeister, Yana Kocheva, Christian Meinhardt, Lukasz Nawacki, Vitor Nunes, Arpad Panyko, Maria Lourdes Ruiz-Rebollo, Cesáreo Flórez-Pardo, Veit Phillip, Aldis Pukitis, Ecaterina Rinja, Vasile Sandru, Arlett Schäfer, Rahel Scholz, Julian Seelig, Simon Sirtl, Diana Vaselane, Volker Ellenrieder, Albrecht Neesse

## Abstract

**Background:** Post-discharge morbidity and mortality is high in acute pancreatitis (AP) and pathophysiological mechanisms remain poorly understood.

**Objectives:** We aim to investigate the composition of gut microbiota and clinical long-term outcomes of prospectively enrolled AP patients to predict post-discharge complications.

**Design:** In this long-term follow-up study, we analysed clinical and microbiome data of 277 patients from the prospective multi-centre P-MAPS trial. Primary endpoint was the association of the microbial composition with post-discharge mortality, recurrent AP (RAP), progression to chronic pancreatitis (CP), pancreatic exocrine insufficiency (PEI), diabetes mellitus (DM) and pancreatic ductal adenocarcinoma (PDAC).

**Results:** Buccal (n=238) and rectal (n=249) swabs were analysed by 16S rRNA and metagenomics sequencing using Oxford Nanopore Technologies. Median follow-up was 2.8 years. Distance-based redundancy analysis (dbRDA) with canonical analysis of principle coordinates (CAP) showed significant differences for β-diversity (Bray-Curtis) for post-discharge mortality (p=0.04), RAP (p=0.02), and DM (p=0.03). A ridge regression model including 11 differentially abundant species predicted post-discharge DM with an area under the receiving operating characteristic (AUROC) of 94.8% and 86.2% in the matched and entire cohort, respectively. Using this classifier, a positive predictive value of 66.6%, a negative predictive value of 96% and an accuracy of 95% was achieved.

**Conclusion:** Our data indicate that the admission microbiome of AP patients correlates with post-discharge complications independent from multiple risk factors such as AP severity, smoking or alcohol. Microbiota at admission show excellent discriminative capacity to predict post-discharge DM and may thus open new stratification tools for a tailored risk assessment in the future.

## Introduction

Acute pancreatitis (AP) is a common inflammatory disease that often leads to hospital admissions. [1] While most patients experience mild or moderately severe disease, 10-20% of patients develop severe inflammation with infected abdominal necrosis, organ failure and increased morbidity and in-hospital mortality. [2] Notably, the clinical management of severe AP has considerably evolved over the last years thereby reducing in-hospital mortality rates by 35%, bringing the overall mortality of AP down to 2-3%. [3] In particular, goal-directed pain and fluid resuscitation approaches, [4, 5] as well as minimally invasive interventions such as endoscopic-ultrasound (EUS) guided drainage of infected necrotic collections have advanced in-hospital management of AP patients. [6] In the past, clinical and scientific interest was primarily focused on these refined minimally-invasive approaches, however, recent data reveal a significant post-discharge morbidity and mortality of AP patients that was long underestimated and understudied. A large prospective, multicentre study from Hungary reported an approximately threefold increased post-acute pancreatitis mortality rate compared to the general population. [7] In line with these observations, two population-based cohort studies from Denmark and Sweden reported similarly increased mortality rates within 90 days post-discharge (around 5%) even exceeding the in-hospital mortality rate. [3, 8] The available literature suggests that post-discharge mortality is higher in patients with more severe AP, and cardiovascular events as well as AP-related sepsis are common causes of death. [3, 7] Furthermore, gastrointestinal diseases including malignancies and cancer-related cachexia are reasons for the increased mortality rate, in particular during the first 2 years after the first episode of AP. [7] While these important clinical studies have prompted follow-up protocols for AP patients after discharge that are increasingly implemented in recent evidence-based guidelines, [9, 10] in particular for patients with moderately severe and severe disease, the exact causes and pathophysiological underpinnings of the increased post-discharge mortality are still poorly understood. It is hypothesized that recurrent acute pancreatitis (RAP), development of chronic pancreatitis (CP), low socioeconomic status and comorbidities considerably contribute to the increased post-discharge mortality. In particular, alcohol-related AP, smoking and male sex are known risk profiles for post-discharge complications. [11] AP-related complications such as the development of diabetes mellitus (DM), pancreatic exocrine insufficiency (PEI), CP and pancreatic ductal adenocarcinoma (PDAC) are likely contributing to the observed post-discharge mortality. [12] Interestingly, a large population-based cohort study showed increased mortality and hospital readmission of post-pancreatitis DM compared to DM type 2 patients, [13] indicating that valid tools for predicting the risk of specific post-discharge complications would greatly assist clinicians to tailor surveillance strategies for AP patients. [12]

We have recently investigated the orointestinal microbiome of AP patients at hospital admission in 15 European centres. Our data revealed striking alterations of microbial patterns in patients with AP that are associated with disease severity and other clinical hallmark features such as length of hospital stay and in-hospital mortality. [14] Here, we hypothesize that the initial admission microbiome of AP patients might be associated with post-discharge mortality, RAP, progression to CP, development of PEI, DM and PDAC over a period of 3 years. For the first time, we construct diagnostic classifiers using differentially abundant species from admission microbiomes that predict post-discharge complications, in particular DM, with high accuracy, linking long-term outcomes of patients with AP to gut microbiota.

## Methods

### Patient cohort and endpoints

Patients with acute pancreatitis from the P-MAPS I cohort (NCT04777812) were contacted between October 2024 and January 2025. [14] Follow-up data were obtained from patientś digital medical records, outpatient visits and phone contact. Local principal investigators collected and stored data pseudonymized in RedCAP. The study endpoints included post-discharge mortality, RAP, progression to CP, development of PEI, DM and PDAC. The P-MAPS I cohort was recruited within 72h after hospital admission in 15 tertiary centres from 8 European countries between March 2020 and June 2022. [15] eSwabs (Copan, Brescia, Italy) were frozen at −80°C within 60 minutes after standardized collection. It was not possible to involve patients or the public in the design, or conduct, or reporting or dissemination plans of our study.

### Microbiome analysis

DNA-extraction, library preparation and sequencing protocols were previously described in detail. [14, 16] In brief, for DNA isolation the PureLink microbiome kit (Invitrogen) was used modified according to International Human Microbiome Standards (IHMS) followed by a purification step with OneStep PCR inhibitor removal kit (Zymo Research). Buccal and rectal samples were sequenced using the whole 16S rRNA and metagenomic approach with Oxford Nanopore Technologies (ONT), respectively. [16] Fast5 files from all samples were re-basecalled using dorado v 1.0.1(ONT). Reads with a mean q-score < 9 were discarded. Subsequently, fastq files were classified using Metapont with a minimum coverage of 50 and alignment score of 1000 with an updated library. A library containing all complete bacterial, fungal, viral as well as mice and human genomes (national center for biotechnology information, NCBI) was used for buccal and rectal samples. The library was established in November 2024.

Microbial analysis was conducted in R 4.4.1 or greater and R-Studio. For normalization purposes, all non-prokaryotic reads were removed and agglomeration to species level was performed. [17] Prevalence filtering with a cut-off at 5% was applied. To control for possible contaminants during sequencing, we analysed 94 blank medium samples (eSwab), pre-PCR water or pre-library preparation water samples undergoing 16S DNA sequencing (n = 38) or metagenomics sequencing (n = 56). Decontam v1.26.0 identified contaminants, which were subsequently removed. Lastly, samples were rarified to 250 000 reads for buccal 16S DNA and to 10 000 reads for rectal metagenomics sequences (Suppl. figure S1).

### Statistical analysis

In the P-MAPS I cohort, 77 discrete and 2 continuous potential confounders with potential impact on microbial composition were prospectively collected. [14] Confounders that occurred at least in 5 % of respective endpoints subgroups were included in a L1-penalized lasso regression, using the respective endpoint as the dependent variable. Afterwards, a tenfold cross-validation was performed to determine the optimal lambda value. Variables with non-zero coefficients in the final model were considered relevant and subsequently included in multiple regression or distance based redundancy analysis of α- and β-diversity calculations.

α-diversity was calculated using observed species (richness) and Shannon index (evenness). Relevant confounders were included in multiple regression models. Given overdispersed count data, a negative binomial regression was fitted for observed species comparison. For Shannon index, a multiple linear regression or multiple gamma regression was utilized depending on normal distribution. β-diversity was determined by Bray-Curtis distances. To stratify for confounders, a distance-based redundancy analysis (db-RDA) ordinated with canonical analysis on the principal coordinates (CAP) was calculated by applying the capscale function of the vegan package (v. 2.6-8). P-values were determined with the permutational ANOVA for β-diversity analysis. 7 values for creatinine and 3 values for body mass index at admission were missing. If one or both of these variables were selected as confounders a multivariate imputation by chained equations was performed (m = 5, mice v. 3.18.0) and the median p-value was displayed. For endpoints with significant different microbial composition in the β-diversity, a microbial classifier was established. To diminish the impact of potential confounders on microbial composition, patients reaching this endpoint were matched against patients who have not met respective endpoints using a random forest approach (MatchIT v. 4.7.1). Differentially abundant species were identified with two different approaches between these matched cohorts: Linear discriminant effect size (LEfSe) and Microbiome Multivariable Association with Linear Models (MaAsLin2). Differentially abundant species were filtered according to their abundances. Here a cut-off between 0.1 % and 0.05 % was chosen to identify 10-15 differentially abundant species. These species were included in a L0-penalized regularisation regression (Ridge). Microbial classifiers were validated with Leave-one-out cross-validation (LOOCV).

## Results

### Patient cohort

In total 424 patients were recruited for the Pancreatitis – Microbiome As Predictor of Severity trial (P-MAPS). From this cohort 10 patients died within 30 days after recruitment. From 120 patients no follow-up data were available. Additionally, 17 patients had a follow-up period of less than 90 days, did not reach an endpoint, and thus were excluded. Out of 277 patients with follow-up data, 181 patients were called by phone, and median follow-up period after initial hospital discharge was 2.8 years (figure 1A, B). Sixty-six (24%) patients RAP, 16 (6.2%) progressed to CP, and 20 (7.2%) died after discharge during the follow-up period. Additionally, 18 (7.3%) developed PEI, 16 (7.2%) were diagnosed DM, and four (1.5%) were diagnosed with PDAC. Among all endpoints, only PEI was associated with severity according to the revised Atlanta classification (RAC, table 1). PDAC was not further evaluated due to the small number of cases.

**Figure 1:**
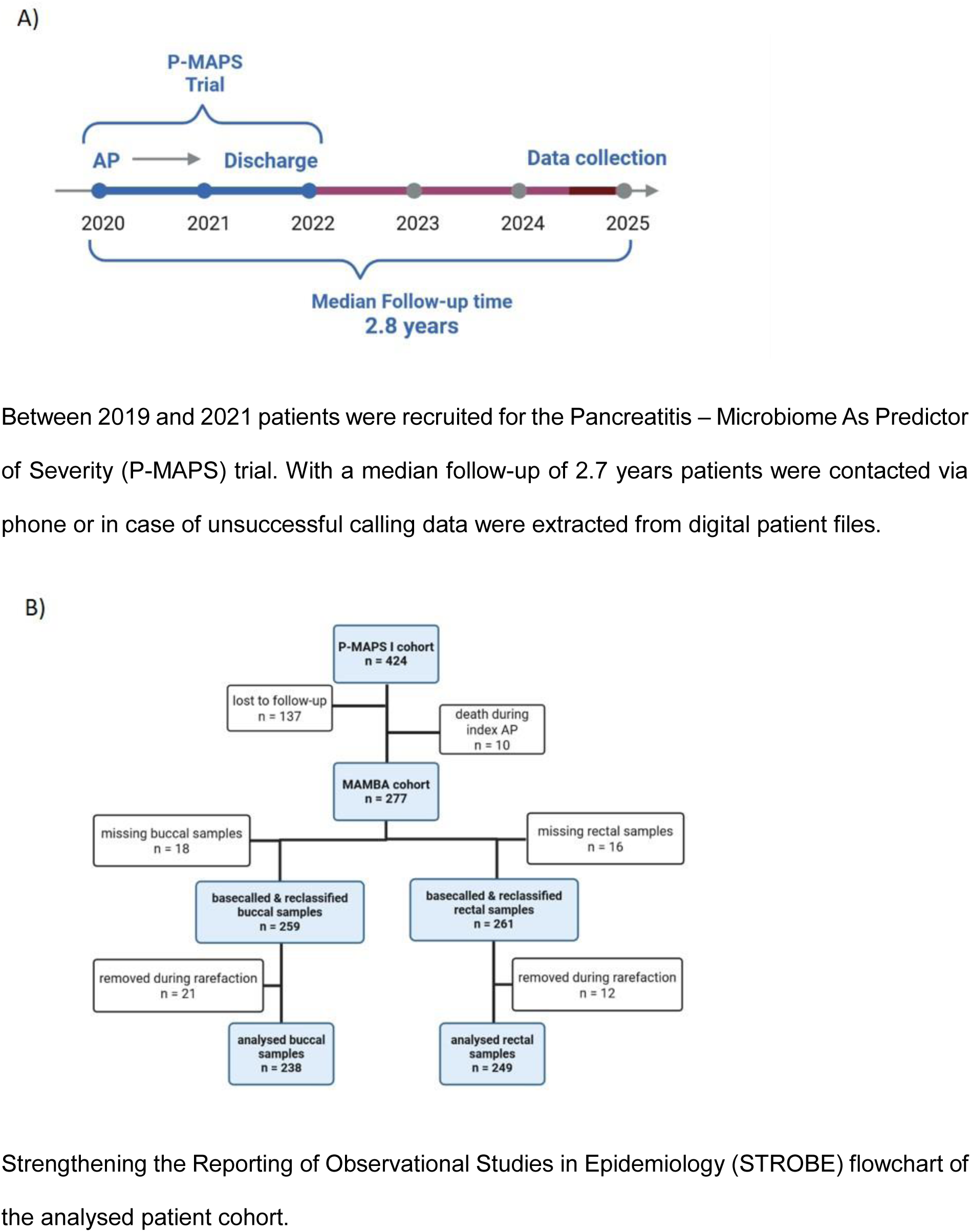
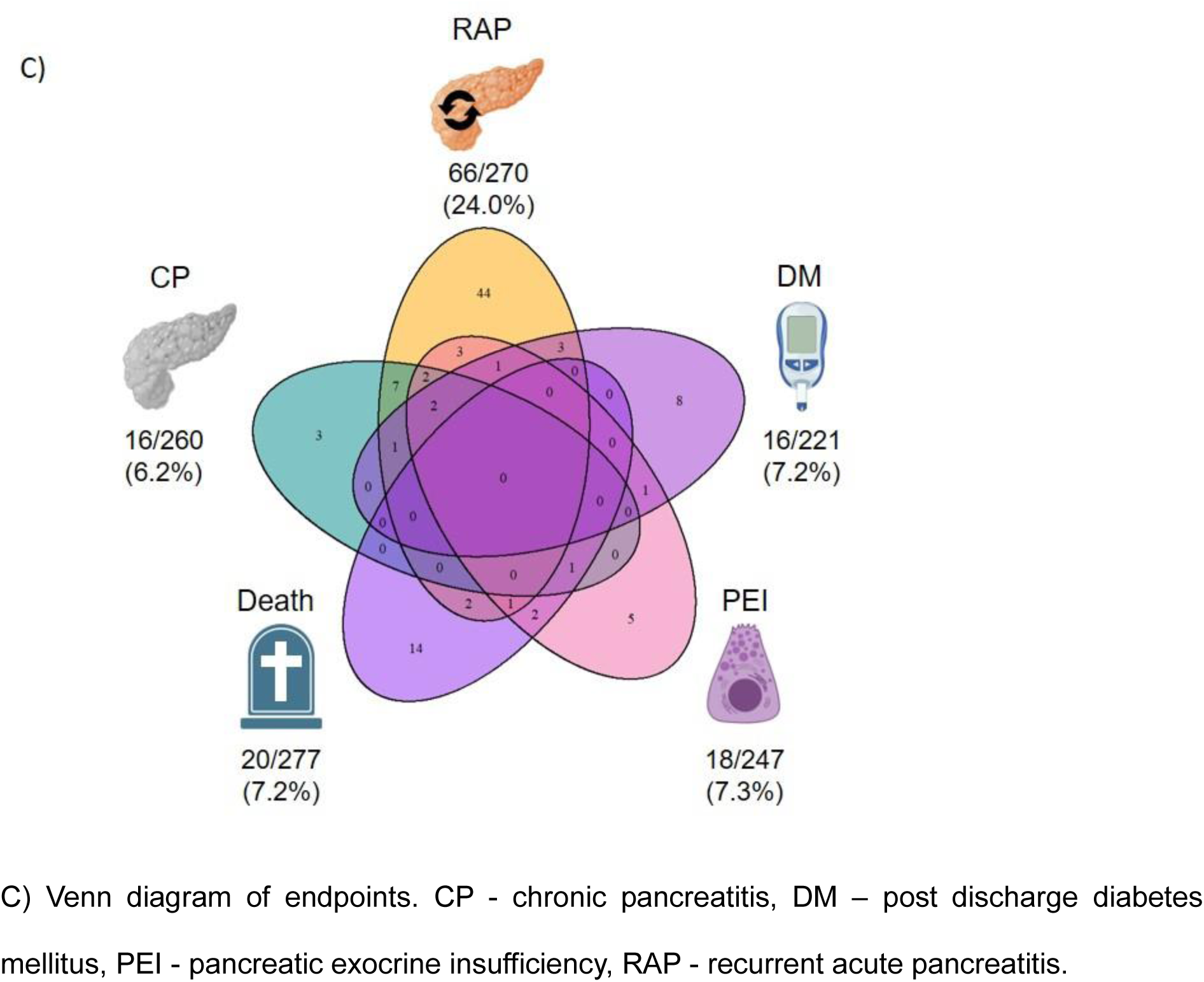
Study overview.

**Table 1:** Endpoints associated with index revised Atlanta classification (RAC) Fisher exact test was performed to reveal possible association between the progression of recurrent acute pancreatitis (RAP), chronic pancreatitis (CP), post discharge mortality, development of post discharge pancreatic exocrine insufficiency (PEI), diabetes and pancreatic ductal adenocarcinoma (PDAC).

| Endpoints | Mild AP<br>(RAC I) | Moderately severe AP<br>(RAC II) | Severe AP<br>(RAC III) | p-value |
| --- | --- | --- | --- | --- |
| **RAP** | 47 / 199<br>(24%) | 17 / 60 (28%) | 2 / 11 (18%) | 0.7 |
| **CP** | 9 / 194<br>(4.6%) | 7 / 55 (13%) | 0 / 11 (0%) | 0.10 |
| **Post discharge**<br>**mortality** | 12 / 202<br>(5.9%) | 7 / 63 (11%) | 1 / 12 (8.3%) | 0.3 |
| **Post discharge PEI** | 8 / 183<br>(4.4%) | 6 / 53 (11%) | 4 / 11 (36%) | **0.001\*\*\*** |
| **Post discharge**<br>**diabetes** | 9 / 165<br>(5.5%) | 5 / 48 (10%) | 2 / 8 (25%) | 0.074 |
| **Post discharge**<br>**PDAC** | 4 / 196<br>(2.0%) | 0 / 57 (0%) | 0 / 11 (2.0%) | 0.6 |

<sup>1</sup>n / N (%)

Of the 277 patients, 259 buccal and 261 rectal microbial data were re-basecalled and reclassified with an updated library. After normalization, 238 buccal and 249 rectal samples had sufficient sequencing depth and were thus eligible for further analysis (figure 1B, Suppl. figure S1). Besides RAP and CP, there was no relevant overlap of patients regarding endpoints (figure 1C). Therefore, associations between the orointestinal microbiome and all endpoints were analysed independently.

### Association between orointestinal microbiome and post-discharge complications Recurrent acute pancreatitis

Out of 238 buccal and 249 rectal samples, five and seven patients had insufficient information regarding recurrent acute pancreatitis (RAP) respectively and were thus excluded for this endpoint. Most patients (49/66) that experienced recurrent acute pancreatitis (RAP) had one or two recurrent episodes after index AP. 11 patients suffered from 3-5 further flares and 6 patients were re-admitted to hospital more than five times due to RAP. As expected, there is a strong association of RAP with smoking and regular alcohol consumption, and an inverse association of biliary cause of AP, cholestasis and older age (Suppl. table 1, 2). Alcohol consumption was determined for stratification of buccal diversity metrics. The evenness was significantly lower in patients with RAP (Shannon: IRR 0.81, p-value 0.015*). No significant differences were obtained for the richness (observed species: incidence rate ration (IRR) 0.97, p-value 0.4) and β-diversity metrics (p-value 0.273, Suppl. figure S2A, B, Suppl. table 3-5). Confounder selection for rectal samples revealed 7 confounding parameters (alcohol consumption, alcoholic AP, cholestasis, opiate intake, moderate and heavy smoking, 1-10 cigarettes/d and > 10 cigarettes/d, and current antibiotic intake) for which multiple models of α- and β-diversity were stratified for. A lower α-diversity was associated with the occurrence of RAP in rectal samples using observed species (IRR 0.94, p-value 0.023*) and the Shannon index (IRR 0.95, p-value 0.056, figure 2A, Suppl. table 6, 7). Notably, significant differences in β-diversity between RAP and non-RAP were revealed in rectal samples using distance-based redundancy analysis (db-RDA, p-value 0.015*, figure 2B, Suppl. table 8).

**Figure 2:**
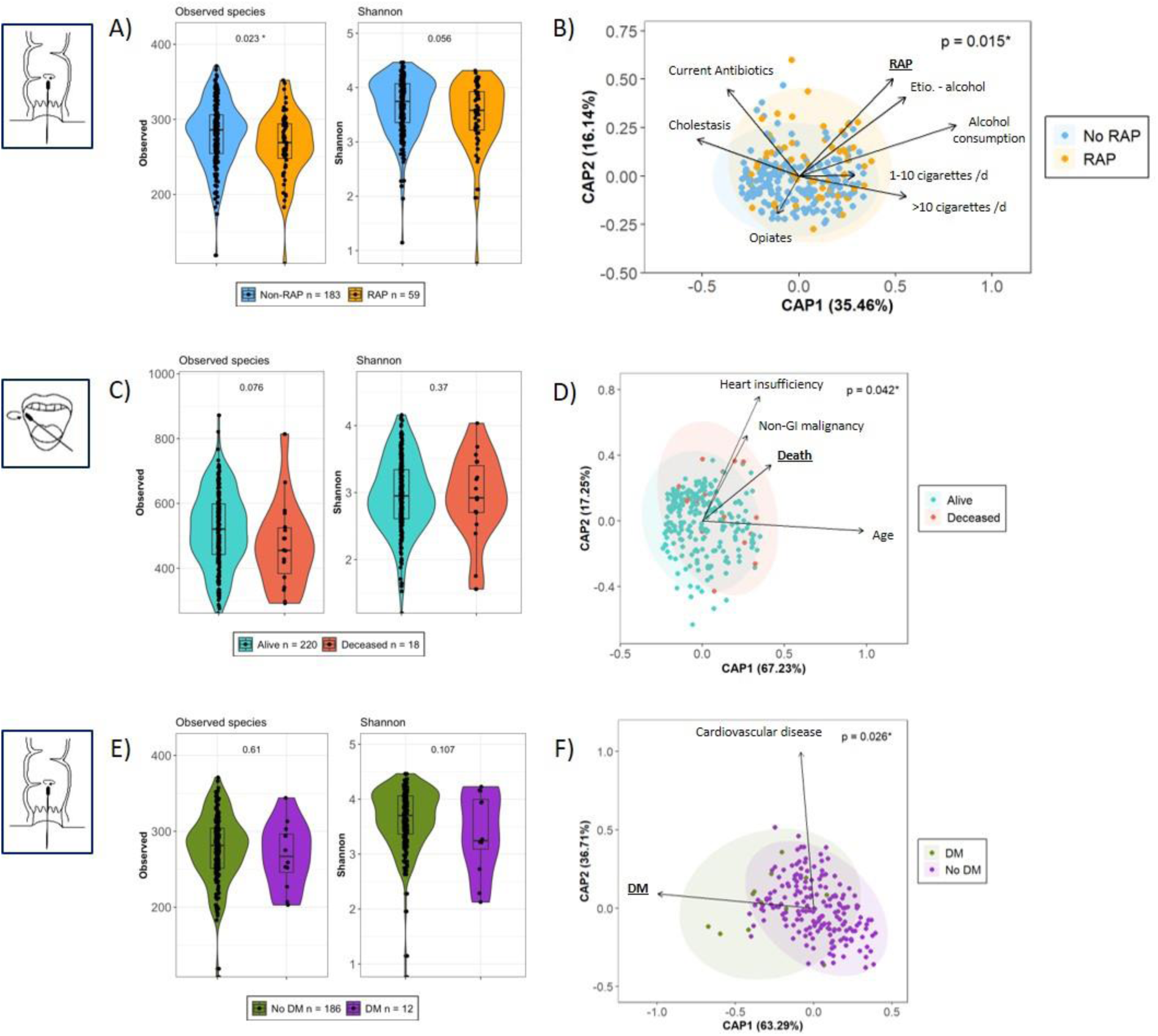
Associations of the admission orointestinal microbiome in acute pancreatitis with post-discharge complications. A) α-diversity was assessed with observed species and Shannon index and was compared between patients with and without a recurrent acute pancreatitis (RAP in orange, Non-RAP in light blue). P-values were calculated using multiple negative binomial regression for observed species and multiple gamma regression for Shannon index in rectal samples. B) Distance-based redundancy analysis was ordinated with canonical analysis of principle coordinates (CAP) between RAP and Non-RAP patients. C) α-diversity metrics for buccal samples were compared between patients died after hospital discharge (violet) and survivors (light green). P-values were calculated using multiple negative binomial regression for observed species and multiple linear regression for Shannon index. D) db-RDA for assessing differences between patients deceased after discharge and survivors. E) α-diversity metrics were compared between patients with and without development of a post-discharge diabetes (No DM in dark green, DM in violet). P-values were calculated using multiple negative binomial regression for observed species and multiple gamma regression for Shannon index in rectal samples. F) Db-RDA for rectal samples differentiates patients with and without the development of post-discharge DM. P-values for db-RDA were calculated using the permutational ANOVA.

### Chronic pancreatitis

Due to unavailable follow-up data, 14 buccal and 17 rectal samples were excluded for α- and β-diversity analysis. 12 out of 16 patients developing CP suffered from prior RAP (figure 1C). Male sex, smoking, current and former alcohol consumption, alcoholic AP and PPI intake were associated with the development of CP (Suppl. table 9, 10). Alcoholic AP, no cholestasis, proton pump inhibitors (PPI) intake and heavy smoking (>10 cigarettes/d) were identified as confounders in both sample sets. Multiple regression models (buccal observed species: IRR 0.99, p-value 0.8; buccal Shannon: IRR 0.75, p-value 0.074; rectal observed species: IRR 1.04, p-value 0.4; rectal Shannon: IRR 1.02, p-value 0.6) and db-RDA (buccal p-value 0.2, rectal p-value 0.6) revealed no significant differences between individuals who developed CP compared to those without CP (Suppl. tables 11-16, Suppl. figure S2C-F).

### Post-discharge mortality

In total 20 patients died within the follow-up period. Half of them died within the first 1.5 years after discharge. The 90-day mortality after discharge was 0.7% (n = 2), and the 12-month mortality was 2.9% (n = 8, Suppl. figure S3). Various causes of death were reported, however, cancer, cardiovascular diseases, sepsis, infections and respiratory diseases were the leading causes (Suppl. table 17). Older age, higher creatinine values at index admission, neurological, malignant and cardiovascular comorbidities and constipation were associated with post-discharge mortality (Suppl. table 18, 19). For buccal samples, confounder selection revealed age, chronic heart insufficiency, and non-gastrointestinal solid malignancies, and all of them were included as co-variates for the α- and β-diversity models. Observed species (IRR 0.90, p-value 0.076) and Shannon index (IRR 0.88, p-value 0.4) were not significantly different between those patients that died after discharge and those that survived (figure 2C, Suppl. table 20, 21), but db-RDA revealed significant differences between both groups for post-discharge mortality (p-value 0.042*, figure 2D, Suppl. table 22). Age, non-gastrointestinal solid malignancy, chronic heart insufficiency and intake of antibiotics within a week before sample collection were considered as confounders for rectal samples. After controlling for confounders, both α-diversity (observed species: IRR 1.03, p-value 0.4; Shannon: IRR 1.01, p-value 0.8) and db-RDA (p-value 0.156) did not show any significant differences (Suppl. figure S2G, H; Suppl. table 23-25).

### Pancreatic exocrine insufficiency

Criteria used for diagnosis of PEI were decreased stool elastase and symptoms of maldigestion. For 26 buccal and 28 rectal samples, corresponding data for PEI were not available and thus excluded from microbial analysis. Alcohol consumption and non-steroidal anti-inflammatory drug (NSAID) intake were associated with the development of PEI (Suppl. table 26, 27). No confounders were identified with L1-penalized lasso regression for buccal and rectal samples, respectively. No significant differences were obtained regarding observed species (buccal: IRR 0.97, p-value 0.6; rectal: IRR 1.03, p-value 0.4), Shannon index (buccal: IRR 0.92, p-value 0.5; rectal Shannon: IRR 1.03, p-value 0.5) and Bray-Curtis distances (buccal: p-value >0.9; rectal: p-value 0.6) in both buccal and rectal samples, respectively (Suppl. figure S2I-L, Suppl. table 28-33).

### Diabetes mellitus

All patients with known DM at index and those without available data for post-discharge DM were excluded. To this end, 47 buccal and 51 rectal samples were not included for analysis. Patients, who developed DM after discharge, differed from patients without DM regarding cardiovascular diseases at index. For buccal samples, statistical tests for α- and β-diversity were not stratified for confounders as L1-penalized lasso regression did not identify variables with non-zero coefficients and yielded no significant alterations between groups (observed species: IRR 0.93, p-value 0.3; Shannon: IRR 0.78, p-value 0.13; Suppl. figure S2M, N, Suppl. table 34-38). Multiple regression models for rectal samples included cardiovascular diseases as covariate and revealed no significant differences in α-diversity for observed species (IRR 0.97, p-value 0.6) and Shannon index (IRR 0.93, p-value 0.11; Suppl. table 39, 40; figure 2E). Remarkably, db-RDA exhibited a significant distance calculation between individuals with and without a development of a post-discharge DM for β-diversity of rectal samples (p value 0.026*, figure 2F, Suppl. table 41).

### Microbial classifier for predicting post-discharge complications

Given the significant alterations of the rectal microbiota associated with the occurrence of RAP, the development of post-discharge DM as well as the associations of changes of the buccal microbiota with post-discharge mortality, we set out to further explore the ability of differentially abundant species to predict these long-term complications with regularization models.

### Recurrent acute pancreatitis

To mitigate confounding effects from clinical variables such as alcohol consumption and smoking, RAP patients were matched to non-RAP controls across confounding variables. This yielded a matched cohort of 53 RAP patients and 81 non-RAP controls for downstream microbial differential abundance analysis (figure 3A, Suppl. Table 42). Using MaAslin2 and LEfSe, we identified 14 species exhibiting significant abundance differences (figure 3B, Suppl. figure S4A, Suppl. table 43). *Finegoldia magna* and *Peptoniphilus harei* were more dominant in the RAP group. Patients without RAP had higher abundances of *Phocaeicola vulgatus* (former *Bacteroides vulgatus*), *Mobiluncus massiliensis* and *Faecalibacterium prausnitzii*. These 14 species were incorporated as predictors in an L0-penalized ridge regression model to classify RAP status. The model demonstrated limited discriminative capacity, achieving an area under the receiver operating characteristics (AUROCs) of 66.5% in the matched cohort and 62.4% in the full cohort (figure 3C).

**Figure 3:**
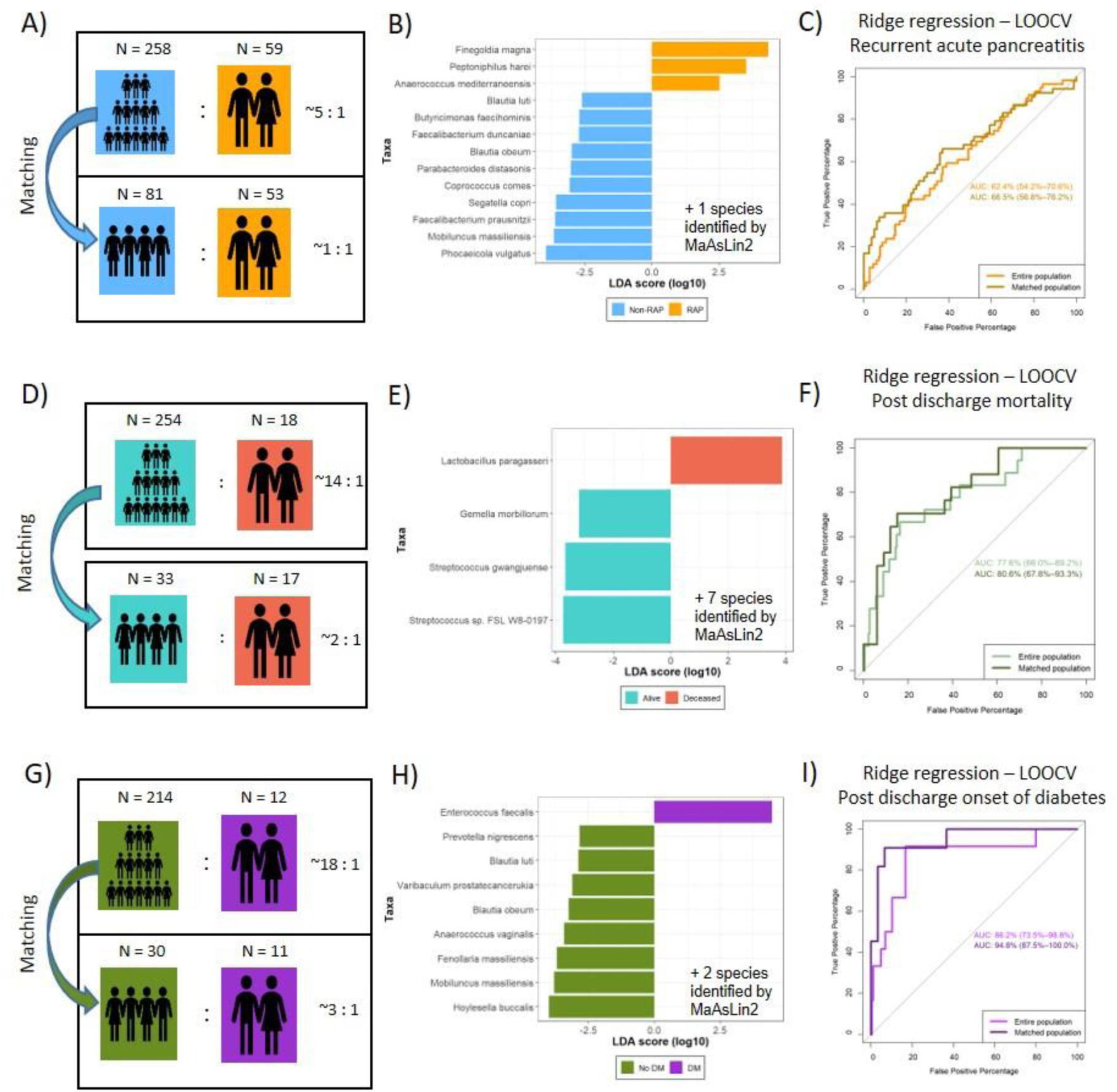
Microbial classifier predicting post-discharge acute pancreatitis related complications. A) Patients with recurrent acute pancreatitis (RAP, orange) were matched with non-RAP controls (light blue). B) Differential abundances of microbial species between groups was assessed using Linear Discriminant Analysis Effect Size (LEfSe). *Anaerococcus prevotii* was additional identified by MaAsLin2. C) An L0-penalized regression model with leave-one-out cross-validation (LOOCV) was applied to differentially abundant species identified by LEfSe and MaAsLin2 in the matched (dark orange) and entire (bright orange) cohorts. D) Patients who died post-discharge (violet) were matched with survivors (light green). E) Differentially abundant species between matched groups were identified using LEfSe. *Terrisporobacter petrolearius, Rummeliibacillus stabekisii, Peptacetobacter hiranonis, Klebsiella electrica, Clostridioides difficile, Exiguobacterium alkaliphilum, Paraclostridium sordellii* were additional identified by MaAsLin2. F) These species, along with those identified by MaAsLin2, were incorporated into an L0-penalized regression model with LOOCV. The area under the receiver operating characteristic curve (AUROC) is shown for the matched (dark green) and entire (light green) cohorts. G) Buccal samples from patients who developed post-discharge diabetes mellitus (DM, violet) and those who did not (non-DM, dark green) were matched. H) LEfSe was used to identify differentially abundant species. *Thomasclavelia ramosa* and *Bacteroides nordii* were additional identified by MaAsLin2. I) An L0-penalized regression classifier was constructed using species identified by LEfSe and MaAsLin2 for the matched (dark violet) and entire (light violet) cohorts.

### Post-discharge mortality

A matched cohort consisting of 33 survivors and 17 patients that died after discharge were identified in a comprehensive matching including potential confounders using random forest (figure 3D Suppl. table 44). Differential abundance revealed 11 species (figure 3E, Suppl. figure S4B, Suppl. table 45). *Lactobacillus paragasseri* was more abundant in patients that died after discharge, whereas *Streptococcus sp. FSL W8-0197* and *Streptococcus gwangjuense* were dominant in the survivor cohort. The performance of the L0-penalized ridge regression was moderate with 80.6% in the matched and 77.6% in the entire cohort, respectively (figure 3F, Suppl. figure 4B).

### Post-discharge diabetes mellitus

Eleven patients, who developed DM after hospital discharge were matched to 30 patients without post-discharge DM (figure 3G, Suppl. table 46). MaAsLin2 and LEfSe calculations identified 11 species that were differentially abundant between both groups. *Enterococcus faecalis* was more dominant in the DM group, whereas *Hoylesella buccalis* and *Mobiluncus massiliensis* had the strongest effect size in the non-DM cohort (figure 3H, Suppl. figure S4C, Suppl. table 47). A ridge regression model including those 11 species was able to predict post-discharge DM with a remarkable AUROC of 94.8% and 86.2% in the matched and entire cohort respectively (figure 3I). These 11 differentially abundant species yielded a positive predictive value of 66.6%, a negative predictive value of 96% and an accuracy of 95%.

## Discussion

Despite improved in-hospital management of AP patients, post-discharge morbidity and mortality remains high, in particular within the first 2 years after the first episode of AP. [7] Increased post-discharged mortality is also known from other conditions such as myocardial infarction or sepsis, [18, 19] highlighting the broader relevance of this topic in clinical medicine. Here, we performed a long-term follow-up study of patients from the prospective, multi-centre P-MAPS trial including 15 centres in 8 European countries. Patients with the first or second episode of AP were enrolled within 72h of hospital admission and oral and rectal microbial swabs were collected according to highly standardized operating procedures. [14–16] Here, we investigated both the frequency of post-discharge mortality and morbidity as well as the potential associations with the admission microbiome following re-classification and re-basecalling with an updated microbiota library. Our clinical data confirm earlier studies revealing a relevant post-discharge mortality of 7.2% within the first 3 years after hospital discharge resulting in a 3-fold increased mortality rate compared to the initial 30 days mortality rate of 2.4% in the P-MAPS cohort. The 90-day mortality (0.7%) was lower in our cohort than in previously reported data (3.0%). [7] Multiple risk factors have been described for the development of post-discharge complications in large population based cohort studies. [11] To this end, our study confirms previous data showing that smoking, continued alcohol abuse, older age, male sex, creatinine at index admission, neurological-, cardiovascular-, and malignant co-morbidities as well as abdominal surgery have a significant impact on the development of post-discharge mortality and related complications. Surprisingly, severity of AP according to RAC was only significantly associated with the development of PEI but no other post-discharge complications. For the subsequent microbiome analysis, we carefully accounted for 77 discrete and 2 continuous potential confounders for microbiome diversity. Distance-based redundancy analysis (db-RDA) ordinated with canonical analysis on the principal coordinates (CAP) clearly showed the pronounced effect of known confounders for post-discharge mortality and RAP such as alcohol consumption, cardiovascular and malignant comorbidities. Despite accounting for these potential confounders in multivariate models, ß-diversity was significantly different for post-discharge mortality, RAP and development of DM indicating a possible link between the microbial composition and these clinical endpoints.

To further reduce the impact of these potential confounders for subsequent construction of diagnostic classifiers, we defined matched cohorts for post-discharge mortality, RAP and DM. The moderate discriminatory capacity of differentially abundant species for post-discharge mortality (77.6%) and RAP (62.4%) can be explained by the relatively strong impact of these confounding factors on both endpoints. In contrast, only previous cardiovascular diseases had to be accounted for in post-discharge development of DM highlighting the strong impact of microbial diversity itself on this clinical endpoint. This strong effect was reflected by a very good discriminatory capacity upon construction of the diagnostic classifier for the development of post-discharge DM (AUROC: 94.8% for the matched cohort, and 86.2% for the entire cohort). Several heterogeneous meta-analyses evaluated the prevalence of DM after one or more episodes of AP and revealed a wide range between 8-54%. [20, 21] Notably, earlier literature on the epidemiology of post-acute pancreatitis DM in population-based studies was prone to selection bias leading to wrong assumptions and incorrect risk constellations. [22] In contrast to large population-based studies, occurrence of post-discharge DM was not significantly associated with AP severity in our cohort pointing towards an important role of the gut-pancreas axis in regulating β-cell function and autoimmunity independent of the extent of pancreas inflammation. However, there is limited knowledge of the pathophysiology of the development of de-novo DM in post-acute pancreatitis. To this end, the gut microbiome might orchestrate a crucial crosstalk between ß-cells and immune cells via antimicrobial peptides. [23] For instance, experimental data in non-obese diabetic (NOD) mice have provided evidence that administration of β-cell produced cathelicidin related antimicrobial peptide (CRAMP) induced regulatory immune cells in pancreatic islets, dampening the incidence of autoimmune diabetes. [24] Intriguingly, CRAMP production by β-cells was controlled by short-chain fatty acids (SCFA) produced by the gut microbiota. Furthermore, dysbiosis in newborn NOD mice induced type 1 interferon production by colonic epithelial cells and promoted subsequent development of pancreatic autoimmune response and development of diabetes providing highlighting an intricate communication between gut microbiota and pancreatic endocrine function. [25] Notably, CRAMP-expressing probiotics restored colonic homeostasis and interrupted the altered glucose metabolism, preventing autoimmune diabetes in NOD mice. [25] These initial findings provide first experimental evidence that microbial treatments using probiotics or even metabolites such as SCFA might hold future promise to prevent post-acute pancreatitis DM.

Interestingly, we found *Prevotella nigrescens*, *Blautia luti*, *Varibaculum protatecancerukia*, *Blautia obeum*, *Anaerococcus vaginalis*, *Fenollaria massiliensus*, *Mobiluncus massiliensis, Thomasclavelia ramosa, Bacteroides nordii* and *Hoylesella buccalis* to be more abundant in the rectal microbiome of patients without post-discharge DM. In contrast, *Enterococcus faecalis* was more abundant in patients with post-discharge DM. Interestingly, *Blautia* was found significantly decreased in patients with new onset DM and restored after adequate antidiabetic medication. [26]

When interpreting our results, several limitations of our study need mentioning. First, the incidence of post-discharge DM was 7.2% and might have been underestimated by the length of follow-up (median 2.8 years) and the lack of standardized HbA1c measurements. However, previous data had shown that the majority of complications occur during the first 2 years after index admission for AP. [7] Second, fecal elastase was not available for most of the patients potentially leading to a possible underestimation of PEI in our study population.

In conclusion, we have conducted a long-term follow-up study of patients with AP showing a striking association of admission gut microbiota and subsequent development of post-discharge complications. Differential abundance analysis and construction of a diagnostic classifier yielded a high discriminatory capacity to predict the development of post-discharge DM thus opening new stratification tools for a tailored risk assessment in the future.

## Data Availability

All data produced in the present study are available upon reasonable request to the authors

## Abbreviations

AUROC: Area under the receiver-operating characteristic
BMI: Body mass index
CAP: Canonical analysis on the principal coordinates
CP: Chronic Pancreatitis
CRAMP: Cathelicidin related antimicrobial peptide
DM: Diabetes Mellitus
Db-RDA: Distance-based redundancy analysis
IRR: Incidence rate ratio
LEfSe: Linear discriminant analysis effect size
LOOCV: Leave-one out cross validation
MaAsLin2: Microbiome Multivariable Association with Linear Models
NCBI: National center for biotechnology information
NOD: Non-obese diabetic (mice)
NSAID: Non-steroidal anti-inflammatory drug
ONT: Oxford-Nanopore Technologies
PDAC: Pancreatic ductal adenocarcinoma
PEI: Pancreatic exocrine insufficiency
P-MAPS: Pancreatitis – Microbiome As Predictor of Severity
PPI: proton pump inhibitors
RAC: Revised Atlanta classification
RAP: Recurrent Acute Pancreatitis
SCFA: Short chain fatty acids

## Acknowledgements

We like to thank Jutta Blumberg and Ulrike Wegner for expert technical assistance. We also thank Alexander Arlt (Oldenburg, Germany), Laura Apadula (Milan, Italy), Georg Beyer (Munich, Germany) and Robert Jaster (Rostock, Germany) for supporting the study at their respective institutions. This study was registered at clinical.trial.gov (NCT04777812).

## Notes

### Competing Interest Statement

The authors have declared no competing interest.

### Clinical Trial

NCT04777812

### Funding Statement

No external funding was received. All sequencing costs were covered by intramural funding of University Medical Center Goettingen, Forschungsfoerderungsprogramm 2021 Startfoerderung Klinische Studien. All authors and institutions received no payment or services from a third party for any aspect of the submitted work.

### Author Declarations

Ethics committee of the University Medical Centre Goettingen, Germany gave ethical approval for this work, protocol number 11/7/19.

