## Supplementary material for "Gut microbiota predict development of post-discharge diabetes mellitus in acute pancreatitis": Suppl Material

**Supplementary files**

Supplementary Figures

A)

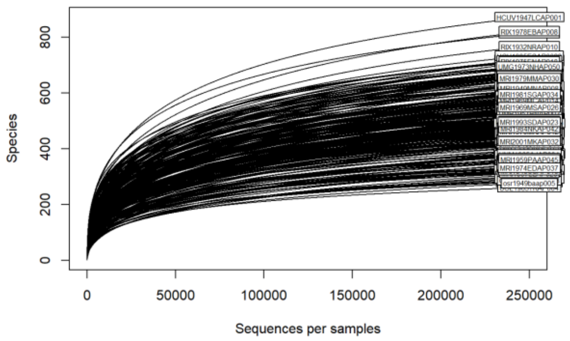


B)

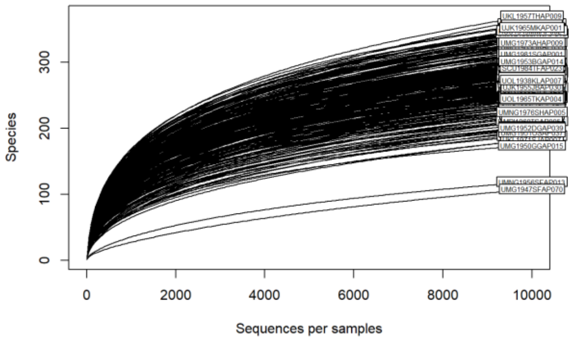


**Suppl. figure S1:** Rarefaction curves
Buccal and rectal samples were rarified with a threshold of 250 000 (16S rRNA sequencing) and 10 000 (metagenomic sequencing) reads per samples, respectively.

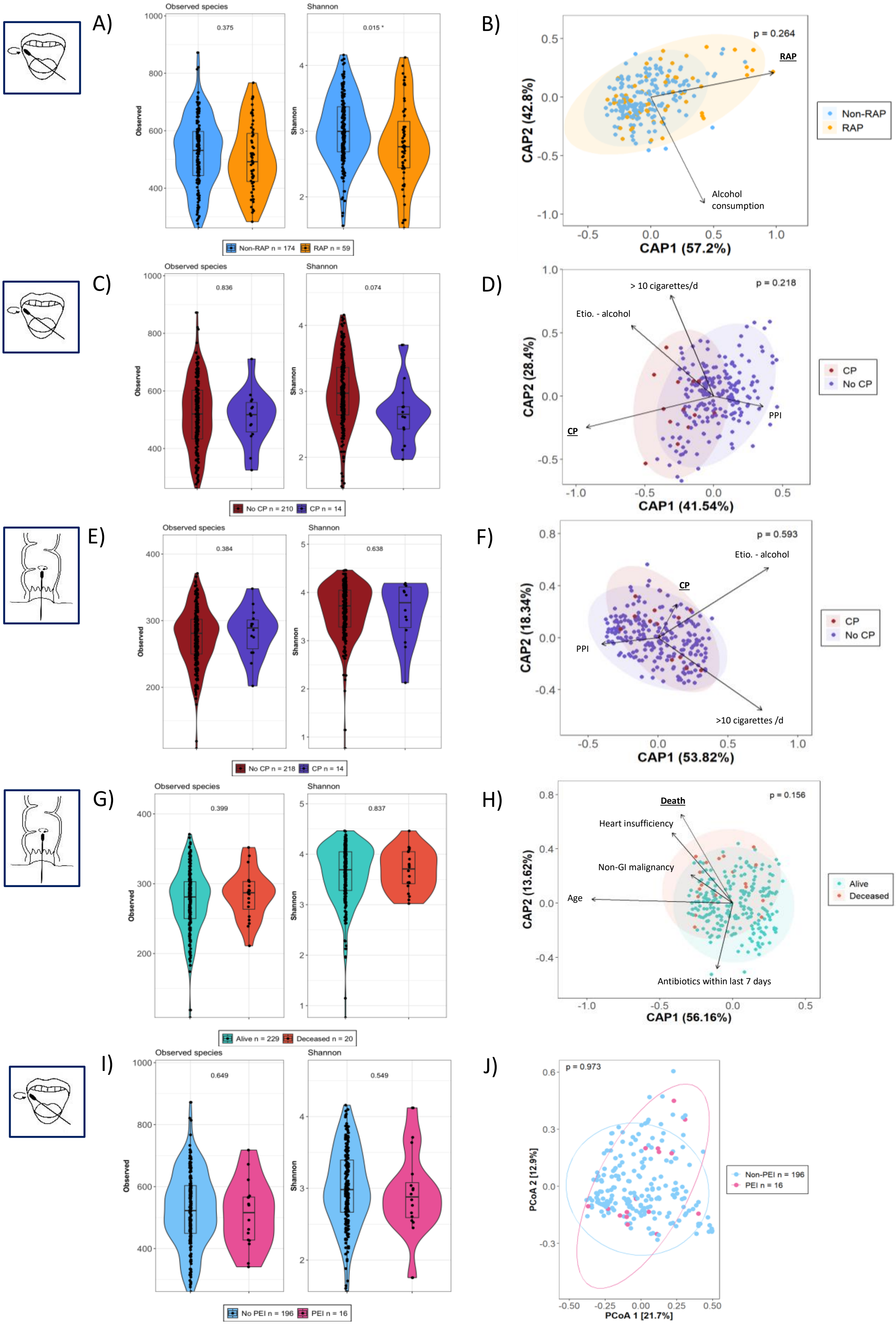
A)
Observed species
Shannon
0.375
0.015 *
Observed
Shannon
Non-RAP n = 174
RAP n = 59
B)
p = 0.264
RAP
Alcohol consumption
CAP2 (42.8%)
CAP1 (57.2%)
Non-RAP
RAP
C)
Observed species
Shannon
0.836
0.074
No CP n = 210
CP n = 14
D)
> 10 cigarettes/d
p = 0.218
Etio. - alcohol
PPI
CP
CAP2 (28.4%)
CAP1 (41.54%)
CP
No CP
E)
Observed species
Shannon
0.384
0.638
No CP n = 218
CP n = 14
F)
p = 0.593
Etio. - alcohol
CP
PPI
>10 cigarettes /d
CAP2 (18.34%)
CAP1 (53.82%)
CP
No CP
G)
Observed species
Shannon
0.399
0.837
Alive n = 229
Deceased n = 20
H)
Death
p = 0.156
Heart insufficiency
Non-GI malignancy
Age
Antibiotics within last 7 days
CAP2 (13.62%)
CAP1 (56.16%)
Alive
Deceased
I)
Observed species
Shannon
0.649
0.549
No PEI n = 196
PEI n = 16
J)
p = 0.973
PCoA 2 [12.9%]
PCoA 1 [21.7%]
Non-PEI n = 196
PEI n = 16

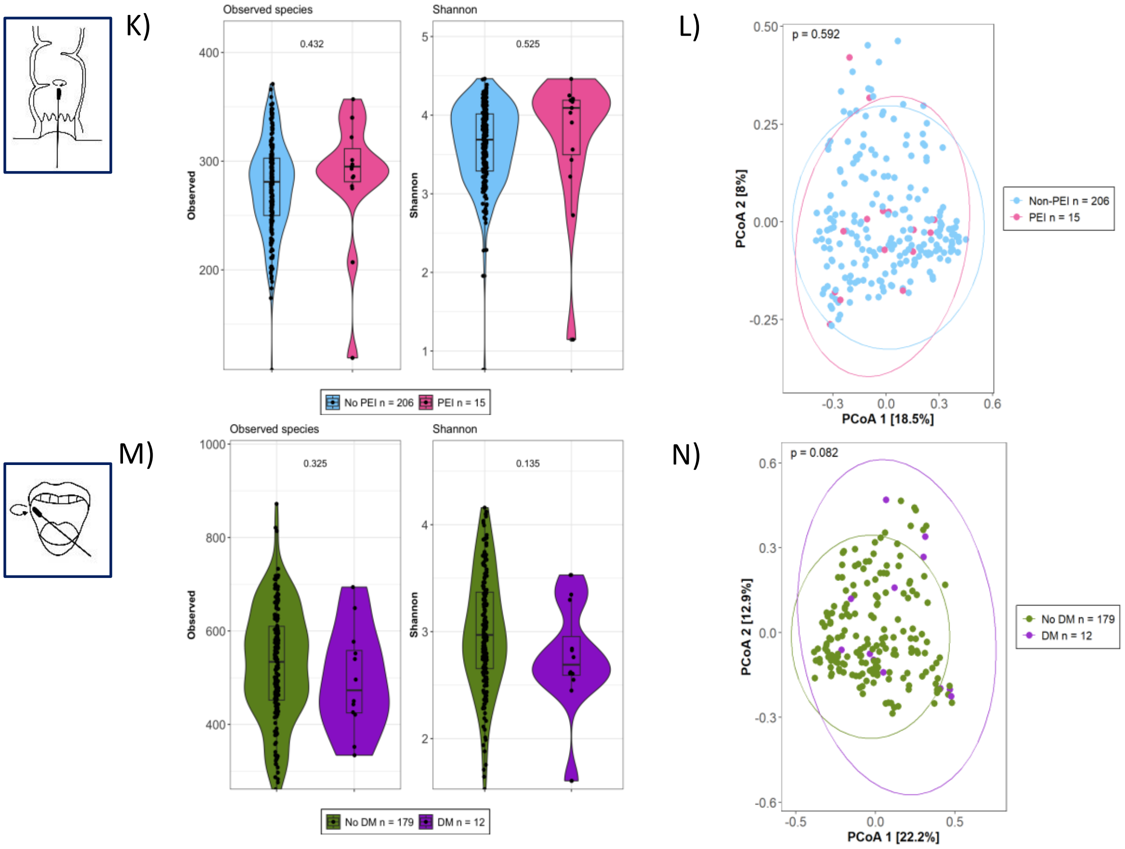


**Suppl. figure S2:** Alpha- and beta diversity for post discharge complications.

A) Observed species and Shannon index of buccal samples was calculated and compared between recurrent acute pancreatitis (RAP, orange) and non-RAP (light blue) patients. B) Distance-based redundancy analysis (db-RDA) of buccal samples between Non-RAP and RAP. C) Alpha diversity metrics of buccal samples from patients with (violet) and without (wine-red) a progression to chronic pancreatitis (CP). D) db-RDA of buccal samples from patients with and without CP. E) Alpha diversity metrics and F) db-RDA of rectal samples comparing CP and Non-CP patients. G) Alpha diversity metrics and H) db-RDA plot of rectal samples from patients who died after hospital discharge (turquoise) and those who were alive in the follow-up period (orange). I) Alpha diversity metrics and J) Bray-Curtis distance of buccal samples from patients with (pink) and without (light blue) the development of a pancreatic exocrine insufficiency (PEI). K) Alpha diversity and L) Bray-Curtis distance of rectal samples from patients with and without the development of PEI. M) Alpha diversity and N) Bray-Curtis distance of buccal samples from patients with (violet) and without (green) the development of a diabetes mellitus (DM). P-values for observed species were calculated using multiple negative binomial regression; for Shannon index, either gamma or linear regression was used based on data normality. db-RDA p-values were derived from permutational ANOVA. GI – Gastrointestinal, NSAID - non-steroidal anti-inflammatory drug

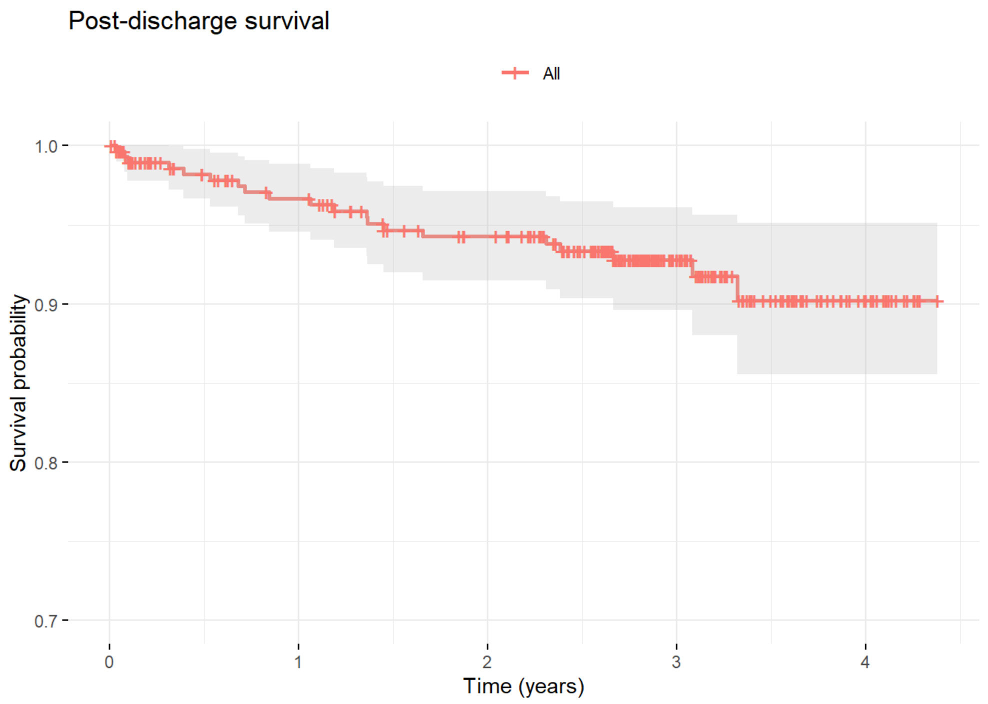


**Suppl. figure S3:** Kaplan-Meyer curve of acute pancreatitis after index discharge

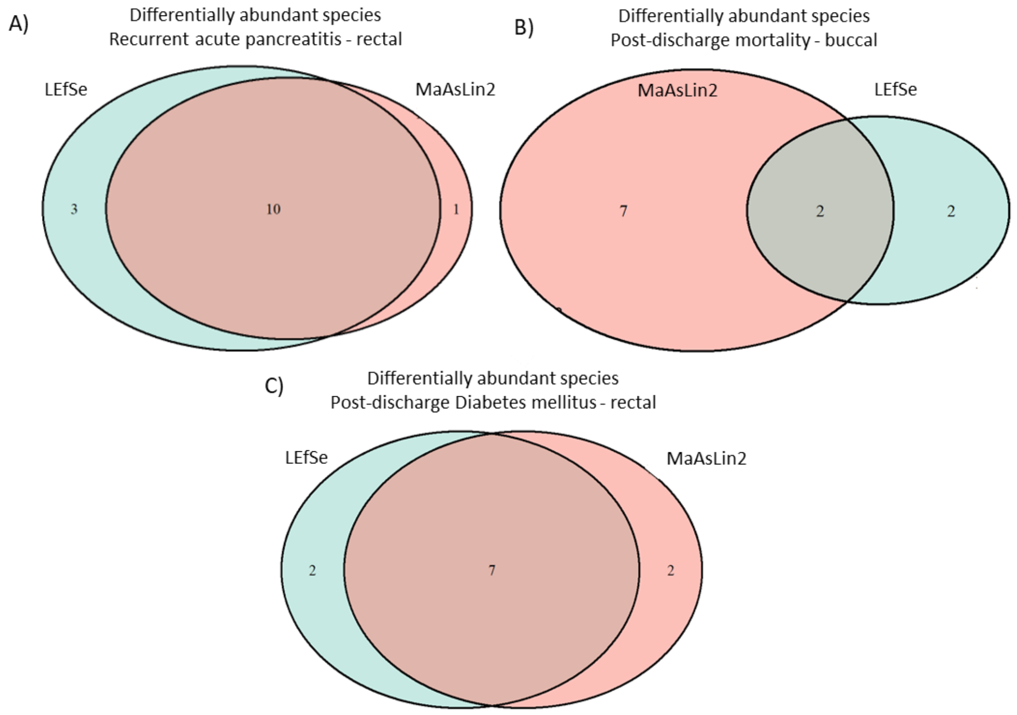


**Suppl. figure S4:** Venn diagram with differentially abundant species.
Species identified as differential abundant for A) recurrent acute pancreatitis, B) post-discharge mortality, and C) post-discharge diabetes mellitus were identified with Linear discriminant analysis effect size (LEfSe, green) and Microbiome Multivariable Association with Linear Models (MaAsLin2, red).

### Supplementary Tables

| **Recurrent acute pancreatitis** | **No** N = 174[1] | **Yes** N = 59[1] | **p-value**[2] |
|---|---|---|---|
| **Gender** | | | 0.3 |
| Female | 80 / 174 (46%) | 23 / 59 (39%) | |
| Male | 94 / 174 (54%) | 36 / 59 (61%) | |
| **Ethnicity** | | | 0.8 |
| African | 1 / 174 (0.6%) | 0 / 59 (0%) | |
| Asian | 1 / 174 (0.6%) | 0 / 59 (0%) | |
| Caucasian | 169 / 174 (97%) | 59 / 59 (100%) | |
| Hispanic or Latino | 3 / 174 (1.7%) | 0 / 59 (0%) | |
| **Etiology** | | | **0.005** |
| Alcoholic | 36 / 174 (21%) | 24 / 59 (41%) | |
| Biliary | 89 / 174 (51%) | 18 / 59 (31%) | |
| Idiopathic | 25 / 174 (14%) | 12 / 59 (20%) | |
| Other | 24 / 174 (14%) | 5 / 59 (8.5%) | |
| **Alcohol** | | | **<0.001** |
| Current chronic alcohol consumption | 39 / 174 (22%) | 27 / 59 (46%) | |
| Former chronic alcohol consumption | 17 / 174 (9.8%) | 8 / 59 (14%) | |
| No alcohol consumption | 118 / 174 (68%) | 24 / 59 (41%) | |
| **Nicotine** | | | **0.011** |
| >10 cig./d | 31 / 174 (18%) | 19 / 59 (32%) | |
| 1-10 cig./d | 10 / 174 (5.7%) | 7 / 59 (12%) | |
| Non-smoker | 133 / 174 (76%) | 33 / 59 (56%) | |
| **Diet** | | | 0.6 |
| Omnivorous | 171 / 174 (98%) | 57 / 59 (97%) | |
| Vegetarian | 3 / 174 (1.7%) | 2 / 59 (3.4%) | |
| **Antibiotics** | | | 0.2 |
| Currently | 25 / 174 (14%) | 3 / 59 (5.1%) | |
| In the last seven days | 3 / 174 (1.7%) | 1 / 59 (1.7%) | |
| In the last six months | 13 / 174 (7.5%) | 5 / 59 (8.5%) | |
| More than six months ago | 133 / 174 (76%) | 50 / 59 (85%) | |
| **Cholestasis at index** | 46 / 174 (26%) | 8 / 59 (14%) | **0.043** |
| **No prior diseases** | 74 / 174 (43%) | 22 / 59 (37%) | 0.5 |
| **Cardiovascular disease** | 45 / 174 (26%) | 17 / 59 (29%) | 0.7 |
| **Heart insufficiency** | 12 / 174 (6.9%) | 6 / 59 (10%) | 0.4 |
| **Diabetes mellitus** | 25 / 174 (14%) | 10 / 59 (17%) | 0.6 |
| **Inflammatory bowel disease** | 4 / 174 (2.3%) | 0 / 59 (0%) | 0.6 |
| **Irritable bowel disease** | 3 / 174 (1.7%) | 1 / 59 (1.7%) | >0.9 |
| **Clostridioides diff. within last 12 month** | | | |
| No | 174 / 174 (100%) | 59 / 59 (100%) | |
| **Chronic constipation** | 4 / 174 (2.3%) | 2 / 59 (3.4%) | 0.6 |
| **Chronic diarrhea** | 1 / 174 (0.6%) | 0 / 59 (0%) | >0.9 |
| **MASLD** | 9 / 174 (5.2%) | 1 / 59 (1.7%) | 0.5 |
| **Liver cirrhosis** | 2 / 174 (1.1%) | 2 / 59 (3.4%) | 0.3 |
| **Other gastrointestinal disease** | 4 / 174 (2.3%) | 2 / 59 (3.4%) | 0.6 |
| **GI cancer** | 5 / 174 (2.9%) | 1 / 59 (1.7%) | >0.9 |
| **Non-GI cancer** | 10 / 174 (5.7%) | 6 / 59 (10%) | 0.2 |

| Recurrent acute pancreatitis | **No** N = 174[1] | **Yes** N = 59[1] | **p-value**[2] |
|---|---|---|---|
| **Hematologic cancer** | 3 / 170 (1.8%) | 0 / 56 (0%) | >0.9 |
| Unknown | 4 | 3 | |
| **Parkinson** | 1 / 174 (0.6%) | 0 / 59 (0%) | >0.9 |
| **Dementia** | 2 / 174 (1.1%) | 1 / 59 (1.7%) | >0.9 |
| **Depression** | 8 / 174 (4.6%) | 2 / 59 (3.4%) | >0.9 |
| **Autism** | | | |
| No | 174 / 174 (100%) | 59 / 59 (100%) | |
| **Other liver disease** | 3 / 174 (1.7%) | 0 / 59 (0%) | 0.6 |
| **GI surgery** | 3 / 174 (1.7%) | 1 / 59 (1.7%) | >0.9 |
| **Abdominal surgery** | 24 / 174 (14%) | 13 / 59 (22%) | 0.13 |
| **HIV** | | | |
| No | 174 / 174 (100%) | 59 / 59 (100%) | |
| **Rheumatic disease** | 9 / 174 (5.2%) | 0 / 59 (0%) | 0.12 |
| **No medication** | 59 / 174 (34%) | 20 / 59 (34%) | >0.9 |
| **Metformin** | 13 / 174 (7.5%) | 7 / 59 (12%) | 0.3 |
| **Other diabetes medication** | 5 / 174 (2.9%) | 4 / 59 (6.8%) | 0.2 |
| **Insulin** | 7 / 174 (4.0%) | 0 / 59 (0%) | 0.2 |
| **Proton-pump inhibitors** | 66 / 174 (38%) | 22 / 59 (37%) | >0.9 |
| **Immunosuppressants** | 7 / 174 (4.0%) | 0 / 59 (0%) | 0.2 |
| **Antidepressants** | 15 / 174 (8.6%) | 2 / 59 (3.4%) | 0.3 |
| **Neuroleptics** | 6 / 174 (3.4%) | 1 / 59 (1.7%) | 0.7 |
| **Paracetamol** | 24 / 174 (14%) | 6 / 59 (10%) | 0.5 |
| **NSAID** | 37 / 174 (21%) | 11 / 59 (19%) | 0.7 |
| **Opiates** | 24 / 174 (14%) | 3 / 59 (5.1%) | 0.071 |
| **Laxatives** | 5 / 174 (2.9%) | 4 / 59 (6.8%) | 0.2 |
| **Statins** | 34 / 174 (20%) | 11 / 59 (19%) | 0.9 |
| **Age** | 59 (18) | 55 (17) | 0.2 |
| **BMI** | 29.0 (5.9) | 27.2 (4.7) | 0.086 |
| Unknown | 6 | 1 | |
| **Admission creatinine (mg/dl)** | 1.00 (0.65) | 1.00 (0.42) | 0.3 |
| Unknown | 3 | 3 | |

[1]n / N (%); Mean (SD)

[2]Pearson's Chi-squared test; Fisher's exact test; Wilcoxon rank sum test

**Suppl. Table 1:** Confounder distribution in buccal samples from patients with recurrent acute pancreatitis (RAP) and non-RAP. Differences were calculated using Fisher´s exact test, Chi-squared test or Mann-Whitney-U test. BMI – body mass index, GI – gastrointestinal, HIV – human immunodeficiency virus, MASLD – metabolic dysfunction-associated steatotic liver disease, NSAID – nonsteroidal anti-inflammatory drug, SD – standard deviation

| Recurrent acute pancreatitis | **No** N = 183[1] | **Yes** N = 59[1] | **p-value**[2] |
|---|---|---|---|
| **Gender** | | | 0.2 |
| Female | 87 / 183 (48%) | 22 / 59 (37%) | |
| Male | 96 / 183 (52%) | 37 / 59 (63%) | |
| **Ethnicity** | | | 0.8 |
| African | 1 / 183 (0.5%) | 0 / 59 (0%) | |
| Asian | 1 / 183 (0.5%) | 0 / 59 (0%) | |
| Caucasian | 177 / 183 (97%) | 59 / 59 (100%) | |
| Hispanic or Latino | 4 / 183 (2.2%) | 0 / 59 (0%) | |
| **Etiology** | | | **<0.001** |
| Alcoholic | 35 / 183 (19%) | 26 / 59 (44%) | |
| Biliary | 100 / 183 (55%) | 17 / 59 (29%) | |
| Idiopathic | 24 / 183 (13%) | 10 / 59 (17%) | |
| Other | 24 / 183 (13%) | 6 / 59 (10%) | |
| **Alcohol** | | | **<0.001** |
| Current chronic alcohol consumption | 39 / 183 (21%) | 26 / 59 (44%) | |
| Former chronic alcohol consumption | 21 / 183 (11%) | 9 / 59 (15%) | |
| No alcohol consumption | 123 / 183 (67%) | 24 / 59 (41%) | |
| **Nicotine** | | | **<0.001** |
| >10 cig./d | 26 / 183 (14%) | 20 / 59 (34%) | |
| 1-10 cig./d | 10 / 183 (5.5%) | 8 / 59 (14%) | |
| Non-smoker | 147 / 183 (80%) | 31 / 59 (53%) | |
| **Diet** | | | 0.6 |
| Omnivorous | 180 / 183 (98%) | 57 / 59 (97%) | |
| Vegetarian | 3 / 183 (1.6%) | 2 / 59 (3.4%) | |
| **Antibiotics** | | | **0.031** |
| Currently | 34 / 183 (19%) | 3 / 59 (5.1%) | |
| In the last seven days | 6 / 183 (3.3%) | 1 / 59 (1.7%) | |
| In the last six months | 18 / 183 (9.8%) | 4 / 59 (6.8%) | |
| More than six months ago | 125 / 183 (68%) | 51 / 59 (86%) | |
| **Cholestasis at index** | 54 / 183 (30%) | 6 / 59 (10%) | **0.003** |
| **No prior diseases** | 70 / 183 (38%) | 22 / 59 (37%) | 0.9 |
| **Cardiovascular disease** | 48 / 183 (26%) | 15 / 59 (25%) | >0.9 |
| **Heart insufficiency** | 14 / 183 (7.7%) | 5 / 59 (8.5%) | 0.8 |
| **Diabetes mellitus** | 26 / 183 (14%) | 10 / 59 (17%) | 0.6 |
| **Inflammatory bowel disease** | 4 / 183 (2.2%) | 0 / 59 (0%) | 0.6 |
| **Irritable bowel disease** | 4 / 183 (2.2%) | 2 / 59 (3.4%) | 0.6 |
| **Clostridioides diff. within last 12 month** | | | |
| No | 183 / 183 (100%) | 59 / 59 (100%) | |
| **Chronic constipation** | 5 / 183 (2.7%) | 2 / 59 (3.4%) | 0.7 |
| **Chronic diarrhea** | 2 / 183 (1.1%) | 1 / 59 (1.7%) | 0.6 |
| **MASLD** | 8 / 183 (4.4%) | 1 / 59 (1.7%) | 0.7 |
| **Liver cirrhosis** | 3 / 183 (1.6%) | 2 / 59 (3.4%) | 0.6 |
| **Other gastrointestinal disease** | 4 / 183 (2.2%) | 3 / 59 (5.1%) | 0.4 |
| **GI malignancy** | 5 / 183 (2.7%) | 1 / 59 (1.7%) | >0.9 |
| **Non-GI malignancy** | 12 / 183 (6.6%) | 6 / 59 (10%) | 0.4 |

| Recurrent acute pancreatitis | **No** N = 183[1] | **Yes** N = 59[1] | p-value[2] |
|---|---|---|---|
| **Hematologic malignancy** | 3 / 181 (1.7%) | 0 / 58 (0%) | >0.9 |
| Unknown | 2 | 1 | |
| **Parkinson** | 3 / 183 (1.6%) | 0 / 59 (0%) | >0.9 |
| **Dementia** | 2 / 183 (1.1%) | 1 / 59 (1.7%) | 0.6 |
| **Depression** | 11 / 183 (6.0%) | 2 / 59 (3.4%) | 0.7 |
| **Autism** | | | |
| No | 183 / 183 (100%) | 59 / 59 (100%) | |
| **Other liver disease** | 4 / 183 (2.2%) | 0 / 59 (0%) | 0.6 |
| **GI surgery** | 4 / 183 (2.2%) | 1 / 59 (1.7%) | >0.9 |
| **Abdominal surgery** | 31 / 183 (17%) | 13 / 59 (22%) | 0.4 |
| **HIV** | | | |
| No | 183 / 183 (100%) | 59 / 59 (100%) | |
| **Rheumatic disease** | 9 / 183 (4.9%) | 0 / 59 (0%) | 0.12 |
| **No medication** | 58 / 183 (32%) | 21 / 59 (36%) | 0.6 |
| **Metformin** | 13 / 183 (7.1%) | 7 / 59 (12%) | 0.3 |
| **Other diabetes medication** | 3 / 183 (1.6%) | 4 / 59 (6.8%) | 0.062 |
| **Insulin** | 7 / 183 (3.8%) | 0 / 59 (0%) | 0.2 |
| **Proton-pump inhibitors** | 78 / 183 (43%) | 22 / 59 (37%) | 0.5 |
| **Immunosuppressants** | 8 / 183 (4.4%) | 0 / 59 (0%) | 0.2 |
| **Antidepressants** | 18 / 183 (9.8%) | 1 / 59 (1.7%) | 0.050 |
| **Neuroleptics** | 5 / 183 (2.7%) | 1 / 59 (1.7%) | >0.9 |
| **Paracetamol** | 26 / 183 (14%) | 7 / 59 (12%) | 0.6 |
| **NSAID** | 41 / 183 (22%) | 13 / 59 (22%) | >0.9 |
| **Opiates** | 29 / 183 (16%) | 4 / 59 (6.8%) | 0.078 |
| **Laxatives** | 5 / 183 (2.7%) | 4 / 59 (6.8%) | 0.2 |
| **Statins** | 39 / 183 (21%) | 10 / 59 (17%) | 0.5 |
| **Age** | 60 (18) | 55 (17) | **0.045** |
| **BMI** | 28.4 (5.5) | 27.6 (4.8) | 0.5 |
| Unknown | 6 | 2 | |
| **Admission creatinine (mg/dl)** | 0.99 (0.65) | 1.00 (0.42) | 0.3 |
| Unknown | 3 | 3 | |

[1]n / N (%); Mean (SD)

[2]Pearson's Chi-squared test; Fisher's exact test; Wilcoxon rank sum test

**Suppl. Table 2:** Confounder distribution in rectal samples from patients with recurrent acute pancreatitis (RAP) and non-RAP. Differences were calculated using Fisher´s exact test, Chi-squared test or Mann-Whitney-U test. BMI – body mass index, GI – gastrointestinal, HIV – human immunodeficiency virus, MASLD – metabolic dysfunction-associated steatotic liver disease, NSAID – nonsteroidal anti-inflammatory drug, SD – standard deviation

| Variables | IRR | 95% CI | p-value |
|---|---|---|---|
| **RAP** | | | |
| *No* | — | — | |
| *Yes* | 0.97 | 0.90, 1.04 | 0.4 |
| **Alcohol consumption** | 0.99 | 0.92, 1.06 | 0.8 |

Abbreviations: CI = Confidence Interval, IRR = Incidence Rate Ratio

**Suppl. Table 3:** Multiple negative binomial regression for observed species in buccal samples. CI – Confidence interval, IRR – incidence rate ratio, RAP – Recurrent acute pancreatitis

| Variables | IRR | 95% CI[1] | p-value |
|---|---|---|---|
| **RAP** | | | |
| *No* | — | — | |
| *Yes* | 0.81 | -0.37, -0.04 | **0.015** |
| **Alcohol consumption** | 0.93 | -0.23, 0.08 | 0.4 |

[1]CI = Confidence Interval

**Suppl. Table 4:** Multiple linear regression for Shannon index in buccal samples.
CI – Confidence interval, Beta – Regression coefficient, RAP – Recurrent acute pancreatitis

| Variables | p-value |
|---|---|
| RAP | 0.264 |
| Alcohol consumption | 0.599 |

**Suppl. Table 5:** Distance-based redundancy analysis (db-RDA) for buccal samples. RAP – Recurrent acute pancreatitis

| Variables | IRR | 95% CI | p-value |
|---|---|---|---|
| **RAP** | | | |
| *No* | — | — | |
| *Yes* | 0.94 | 0.90, 0.99 | **0.023** |
| **Opiates** | | | |
| *No* | — | — | |
| *Yes* | 0.98 | 0.92, 1.04 | 0.5 |
| **Alcohol consumption** | 1.01 | 0.95, 1.08 | 0.8 |
| **Current antibiotics** | 0.94 | 0.89, 1.00 | **0.047** |
| **Etiology - Alcohol** | 0.96 | 0.90, 1.03 | 0.3 |
| **Nicotine consumption** | | | |
| *>10 cigarettes/d* | — | — | |
| *1-10 cigarettes/d* | 0.99 | 0.91, 1.08 | 0.9 |
| *Non-smoker* | 0.97 | 0.91, 1.02 | 0.2 |
| **Cholestasis** | | | |
| *No* | — | — | |
| *Yes* | 1.04 | 0.99, 1.09 | 0.13 |

Abbreviations: CI = Confidence Interval, IRR = Incidence Rate Ratio

**Suppl. Table 6:** Multiple negative binomial regression for observed species in rectal samples. CI – Confidence interval, IRR – incidence rate ratio, RAP – Recurrent acute pancreatitis

| Variables | IRR | 95% CI | p-value |
|---|---|---|---|
| **RAP** | | | |
| *No* | — | — | |
| *Yes* | 0.95 | 0.91, 1.00 | 0.056 |
| **Opiates** | | | |
| *No* | — | — | |
| *Yes* | 1.01 | 0.96, 1.08 | 0.6 |
| **Alcohol consumption** | 0.99 | 0.93, 1.06 | 0.9 |
| **Current antibiotics** | 0.94 | 0.89, 1.00 | **0.043** |
| **Etiology - Alcohol** | 0.97 | 0.90, 1.04 | 0.4 |
| **Nicotine consumption** | | | |
| *>10 cigarettes/d* | — | — | |
| *1-10 cigarettes/d* | 0.98 | 0.90, 1.07 | 0.6 |
| *Non-smoker* | 0.98 | 0.93, 1.04 | 0.5 |
| **Cholestasis** | | | |
| *No* | — | — | |
| *Yes* | 1.00 | 0.96, 1.06 | 0.8 |

Abbreviation: CI = Confidence Interval

**Suppl. Table 7:** Multiple gamma regression for Shannon index in rectal samples.
CI – Confidence interval, IRR – incidence rate ratio, RAP – Recurrent acute pancreatitis

| Variables | p-value |
|---|---|
| RAP | **0.015*** |
| Nicotine consumption | 0.429 |

| | |
|---|---|
| Current antibiotics | 0.163 |
| Alcohol consumption | **0.002**** |
| Cholestasis at index | 0.197 |
| Opiates | 0.94 |
| Etiology - Alcohol | 0.216 |

**Suppl. Table 8:** Distance-based redundancy analysis (db-RDA) for rectal samples. RAP – Recurrent acute pancreatitis

| **Chronic pancreatitis** | **No** N = 210[1] | **Yes** N = 14[1] | **p-value**[2] |
|---|---|---|---|
| **Gender** | | | **0.028** |
| Female | 93 / 210 (44%) | 2 / 14 (14%) | |
| Male | 117 / 210 (56%) | 12 / 14 (86%) | |
| **Ethnicity** | | | >0.9 |
| African | 1 / 210 (0.5%) | 0 / 14 (0%) | |
| Asian | 1 / 210 (0.5%) | 0 / 14 (0%) | |
| Caucasian | 205 / 210 (98%) | 14 / 14 (100%) | |
| Hispanic or Latino | 3 / 210 (1.4%) | 0 / 14 (0%) | |
| **Etiology** | | | **<0.001** |
| Alcoholic | 47 / 210 (22%) | 11 / 14 (79%) | |
| Biliary | 101 / 210 (48%) | 2 / 14 (14%) | |
| Idiopathic | 35 / 210 (17%) | 0 / 14 (0%) | |
| Other | 27 / 210 (13%) | 1 / 14 (7.1%) | |
| **Alcohol** | | | **0.004** |
| Current chronic alcohol consumption | 55 / 210 (26%) | 9 / 14 (64%) | |
| Former chronic alcohol consumption | 22 / 210 (10%) | 2 / 14 (14%) | |
| No alcohol consumption | 133 / 210 (63%) | 3 / 14 (21%) | |
| **Nicotine** | | | **0.004** |
| >10 cig./d | 41 / 210 (20%) | 8 / 14 (57%) | |
| 1-10 cig./d | 15 / 210 (7.1%) | 1 / 14 (7.1%) | |
| Non-smoker | 154 / 210 (73%) | 5 / 14 (36%) | |
| **Diet** | | | >0.9 |
| Omnivorous | 206 / 210 (98%) | 14 / 14 (100%) | |
| Vegetarian | 4 / 210 (1.9%) | 0 / 14 (0%) | |
| **Antibiotics** | | | 0.4 |
| Currently | 26 / 210 (12%) | 0 / 14 (0%) | |
| In the last seven days | 4 / 210 (1.9%) | 0 / 14 (0%) | |
| In the last six months | 17 / 210 (8.1%) | 0 / 14 (0%) | |
| More than six months ago | 163 / 210 (78%) | 14 / 14 (100%) | |
| **Cholestasis at index** | 52 / 210 (25%) | 0 / 14 (0%) | **0.044** |
| **No prior diseases** | 87 / 210 (41%) | 5 / 14 (36%) | 0.7 |
| **Cardiovascular disease** | 58 / 210 (28%) | 3 / 14 (21%) | 0.8 |
| **Heart insufficiency** | 17 / 210 (8.1%) | 0 / 14 (0%) | 0.6 |
| **Diabetes mellitus** | 32 / 210 (15%) | 3 / 14 (21%) | 0.5 |
| **Inflammatory bowel disease** | 4 / 210 (1.9%) | 0 / 14 (0%) | >0.9 |
| **Irritable bowel disease** | 4 / 210 (1.9%) | 0 / 14 (0%) | >0.9 |
| **Clostridioides diff. within last 12 month** | | | |
| No | 210 / 210 (100%) | 14 / 14 (100%) | |
| **Chronic constipation** | 5 / 210 (2.4%) | 0 / 14 (0%) | >0.9 |
| **Chronic diarrhea** | 1 / 210 (0.5%) | 0 / 14 (0%) | >0.9 |
| **MASLD** | 10 / 210 (4.8%) | 0 / 14 (0%) | >0.9 |
| **Liver cirrhosis** | 3 / 210 (1.4%) | 1 / 14 (7.1%) | 0.2 |
| **Other gastrointestinal disease** | 6 / 210 (2.9%) | 0 / 14 (0%) | >0.9 |
| **GI cancer** | 6 / 210 (2.9%) | 0 / 14 (0%) | >0.9 |
| **Non-GI cancer** | 13 / 210 (6.2%) | 2 / 14 (14%) | 0.2 |

| Chronic pancreatitis | **No** N = 210[1] | **Yes** N = 14[1] | p-value[2] |
|---|---|---|---|
| **Hematologic cancer** | 3 / 205 (1.5%) | 0 / 13 (0%) | >0.9 |
| Unknown | 5 | 1 | |
| **Parkinson** | 1 / 210 (0.5%) | 0 / 14 (0%) | >0.9 |
| **Dementia** | 3 / 210 (1.4%) | 0 / 14 (0%) | >0.9 |
| **Depression** | 9 / 210 (4.3%) | 1 / 14 (7.1%) | 0.5 |
| **Autism** | | | |
| No | 210 / 210 (100%) | 14 / 14 (100%) | |
| **Other liver disease** | 3 / 210 (1.4%) | 0 / 14 (0%) | >0.9 |
| **GI surgery** | 4 / 210 (1.9%) | 0 / 14 (0%) | >0.9 |
| **Abdominal surgery** | 33 / 210 (16%) | 2 / 14 (14%) | >0.9 |
| **HIV** | | | |
| No | 210 / 210 (100%) | 14 / 14 (100%) | |
| **Rheumatic disease** | 9 / 210 (4.3%) | 0 / 14 (0%) | >0.9 |
| **No medication** | 71 / 210 (34%) | 6 / 14 (43%) | 0.6 |
| **Metformin** | 18 / 210 (8.6%) | 2 / 14 (14%) | 0.4 |
| **Other diabetes medication** | 9 / 210 (4.3%) | 0 / 14 (0%) | >0.9 |
| **Insulin** | 7 / 210 (3.3%) | 0 / 14 (0%) | >0.9 |
| **Proton-pump inhibitors** | 82 / 210 (39%) | 1 / 14 (7.1%) | **0.017** |
| **Immunosuppressants** | 7 / 210 (3.3%) | 0 / 14 (0%) | >0.9 |
| **Antidepressants** | 16 / 210 (7.6%) | 1 / 14 (7.1%) | >0.9 |
| **Neuroleptics** | 7 / 210 (3.3%) | 0 / 14 (0%) | >0.9 |
| **Paracetamol** | 28 / 210 (13%) | 1 / 14 (7.1%) | >0.9 |
| **NSAID** | 42 / 210 (20%) | 2 / 14 (14%) | >0.9 |
| **Opiates** | 25 / 210 (12%) | 2 / 14 (14%) | 0.7 |
| **Laxatives** | 7 / 210 (3.3%) | 0 / 14 (0%) | >0.9 |
| **Statins** | 43 / 210 (20%) | 1 / 14 (7.1%) | 0.3 |
| **Age** | 58 (18) | 50 (12) | 0.056 |
| **BMI** | 28.7 (5.8) | 27.4 (4.6) | 0.5 |
| Unknown | 7 | 0 | |
| **Admission creatinine (mg/dl)** | 1.00 (0.61) | 0.96 (0.47) | 0.7 |
| Unknown | 4 | 0 | |

[1]n / N (%); Mean (SD)

[2]Pearson's Chi-squared test; Fisher's exact test; Wilcoxon rank sum test

**Suppl. Table 9:** Confounder distribution in buccal samples from patients with chronic pancreatitis (CP) and non-CP. Differences were calculated using Fisher´s exact test, Chi-squared test or Mann-Whitney-U test. BMI – body mass index, GI – gastrointestinal, HIV – human immunodeficiency virus, MASLD – metabolic dysfunction-associated steatotic liver disease, NSAID – nonsteroidal anti-inflammatory drug, SD – standard deviation

| Chronic pancreatitis | **No** N = 218[1] | **Yes** N = 14[1] | **p-value**[2] |
|---|---|---|---|
| **Gender** | | | **0.023** |
| Female | 99 / 218 (45%) | 2 / 14 (14%) | |
| Male | 119 / 218 (55%) | 12 / 14 (86%) | |
| **Ethnicity** | | | >0.9 |
| African | 1 / 218 (0.5%) | 0 / 14 (0%) | |
| Asian | 1 / 218 (0.5%) | 0 / 14 (0%) | |
| Caucasian | 212 / 218 (97%) | 14 / 14 (100%) | |
| Hispanic or Latino | 4 / 218 (1.8%) | 0 / 14 (0%) | |
| **Etiology** | | | **<0.001** |
| Alcoholic | 47 / 218 (22%) | 11 / 14 (79%) | |
| Biliary | 111 / 218 (51%) | 2 / 14 (14%) | |
| Idiopathic | 32 / 218 (15%) | 0 / 14 (0%) | |
| Other | 28 / 218 (13%) | 1 / 14 (7.1%) | |
| **Alcohol** | | | **0.014** |
| Current chronic alcohol consumption | 54 / 218 (25%) | 8 / 14 (57%) | |
| Former chronic alcohol consumption | 27 / 218 (12%) | 2 / 14 (14%) | |
| No alcohol consumption | 137 / 218 (63%) | 4 / 14 (29%) | |
| **Nicotine** | | | **0.002** |
| >10 cig./d | 37 / 218 (17%) | 8 / 14 (57%) | |
| 1-10 cig./d | 15 / 218 (6.9%) | 1 / 14 (7.1%) | |
| Non-smoker | 166 / 218 (76%) | 5 / 14 (36%) | |
| **Diet** | | | >0.9 |
| Omnivorous | 214 / 218 (98%) | 14 / 14 (100%) | |
| Vegetarian | 4 / 218 (1.8%) | 0 / 14 (0%) | |
| **Antibiotics** | | | 0.2 |
| Currently | 35 / 218 (16%) | 0 / 14 (0%) | |
| In the last seven days | 7 / 218 (3.2%) | 0 / 14 (0%) | |
| In the last six months | 21 / 218 (9.6%) | 0 / 14 (0%) | |
| More than six months ago | 155 / 218 (71%) | 14 / 14 (100%) | |
| **Cholestasis at index** | 58 / 218 (27%) | 0 / 14 (0%) | **0.024** |
| **No prior diseases** | 83 / 218 (38%) | 5 / 14 (36%) | 0.9 |
| **Cardiovascular disease** | 59 / 218 (27%) | 3 / 14 (21%) | 0.8 |
| **Heart insufficiency** | 18 / 218 (8.3%) | 0 / 14 (0%) | 0.6 |
| **Diabetes mellitus** | 33 / 218 (15%) | 3 / 14 (21%) | 0.5 |
| **Inflammatory bowel disease** | 4 / 218 (1.8%) | 0 / 14 (0%) | >0.9 |
| **Irritable bowel disease** | 5 / 218 (2.3%) | 1 / 14 (7.1%) | 0.3 |
| **Clostridioides diff. within last 12 month** | | | |
| No | 218 / 218 (100%) | 14 / 14 (100%) | |
| **Chronic constipation** | 6 / 218 (2.8%) | 0 / 14 (0%) | >0.9 |
| **Chronic diarrhea** | 2 / 218 (0.9%) | 1 / 14 (7.1%) | 0.2 |
| **MASLD** | 9 / 218 (4.1%) | 0 / 14 (0%) | >0.9 |
| **Liver cirrhosis** | 4 / 218 (1.8%) | 1 / 14 (7.1%) | 0.3 |
| **Other gastrointestinal disease** | 6 / 218 (2.8%) | 0 / 14 (0%) | >0.9 |
| **GI malignancy** | 6 / 218 (2.8%) | 0 / 14 (0%) | >0.9 |
| **Non-GI malignancy** | 15 / 218 (6.9%) | 2 / 14 (14%) | 0.3 |

| Chronic pancreatitis | **No** N = 218[1] | **Yes** N = 14[1] | **p-value**[2] |
|---|---|---|---|
| **Hematologic malignancy** | 3 / 216 (1.4%) | 0 / 14 (0%) | >0.9 |
| Unknown | 2 | 0 | |
| **Parkinson** | 3 / 218 (1.4%) | 0 / 14 (0%) | >0.9 |
| **Dementia** | 3 / 218 (1.4%) | 0 / 14 (0%) | >0.9 |
| **Depression** | 13 / 218 (6.0%) | 0 / 14 (0%) | >0.9 |
| **Autism** | | | |
| No | 218 / 218 (100%) | 14 / 14 (100%) | |
| **Other liver disease** | 4 / 218 (1.8%) | 0 / 14 (0%) | >0.9 |
| **GI surgery** | 5 / 218 (2.3%) | 0 / 14 (0%) | >0.9 |
| **Abdominal surgery** | 40 / 218 (18%) | 2 / 14 (14%) | >0.9 |
| **HIV** | | | |
| No | 218 / 218 (100%) | 14 / 14 (100%) | |
| **Rheumatic disease** | 9 / 218 (4.1%) | 0 / 14 (0%) | >0.9 |
| **No medication** | 70 / 218 (32%) | 6 / 14 (43%) | 0.4 |
| **Metformin** | 18 / 218 (8.3%) | 2 / 14 (14%) | 0.3 |
| **Other diabetes medication** | 7 / 218 (3.2%) | 0 / 14 (0%) | >0.9 |
| **Insulin** | 7 / 218 (3.2%) | 0 / 14 (0%) | >0.9 |
| **Proton-pump inhibitors** | 93 / 218 (43%) | 2 / 14 (14%) | **0.036** |
| **Immunosuppressants** | 8 / 218 (3.7%) | 0 / 14 (0%) | >0.9 |
| **Antidepressants** | 19 / 218 (8.7%) | 0 / 14 (0%) | 0.6 |
| **Neuroleptics** | 6 / 218 (2.8%) | 0 / 14 (0%) | >0.9 |
| **Paracetamol** | 31 / 218 (14%) | 1 / 14 (7.1%) | 0.7 |
| **NSAID** | 47 / 218 (22%) | 3 / 14 (21%) | >0.9 |
| **Opiates** | 30 / 218 (14%) | 3 / 14 (21%) | 0.4 |
| **Laxatives** | 7 / 218 (3.2%) | 0 / 14 (0%) | >0.9 |
| **Statins** | 47 / 218 (22%) | 1 / 14 (7.1%) | 0.3 |
| **Age** | 59 (18) | 51 (12) | 0.055 |
| **BMI** | 28.4 (5.4) | 27.2 (4.8) | 0.4 |
| Unknown | 7 | 0 | |
| **Admission creatinine (mg/dl)** | 0.99 (0.61) | 0.94 (0.47) | 0.6 |
| Unknown | 4 | 0 | |

[1]n / N (%); Mean (SD)

[2]Pearson's Chi-squared test; Fisher's exact test; Wilcoxon rank sum test

**Suppl. Table 10:** Confounder distribution in rectal samples from patients with chronic pancreatitis (CP) and non-CP. Differences were calculated using Fisher´s exact test, Chi-squared test or Mann-Whitney-U test. BMI – body mass index, GI – gastrointestinal, HIV – human immunodeficiency virus, MASLD – metabolic dysfunction-associated steatotic liver disease, NSAID – nonsteroidal anti-inflammatory drug, SD – standard deviation

| Variables | IRR | 95% CI | p-value |
|---|---|---|---|
| **Chronic pancreatitis** | | | |
| *No* | — | — | |
| *Yes* | 0.99 | 0.86, 1.13 | 0.8 |
| **Etiology - Alcohol** | 0.98 | 0.90, 1.06 | 0.6 |
| **> 10 cigarettes / day** | 1.01 | 0.94, 1.10 | 0.7 |
| **Proton-pump inhibitors** | | | |
| *No* | — | — | |
| *Yes* | 1.00 | 0.94, 1.07 | >0.9 |

Abbreviations: CI = Confidence Interval, IRR = Incidence Rate Ratio

**Suppl. Table 11:** Multiple negative binomial regression for observed species in buccal samples. CI – Confidence interval, IRR – incidence rate ratio

| Variables | IRR | 95% CI[1] | p-value |
|---|---|---|---|
| **Chronic pancreatitis** | | | |
| *No* | — | — | |
| *Yes* | 0.75 | -0.60, 0.03 | 0.074 |
| **Etiology - Alcohol** | 0.94 | -0.24, 0.12 | 0.5 |
| **No Nicotine** | 0.97 | -0.21, 0.16 | 0.8 |
| **Proton-pump inhibitors** | | | |
| *No* | — | — | |
| *Yes* | 1.02 | -0.13, 0.17 | 0.8 |

[1]CI = Confidence Interval

**Suppl. Table 12:** Multiple linear regression for Shannon index in buccal samples. CI – Confidence interval, Beta – regression coefficient

| Variables | p-value |
|---|---|
| CP | 0.218 |
| Etio. - Alcohol | 0.787 |
| Proton-pump inhibitors | 0.979 |
| > 10 cigarettes / day | 0.855 |

**Suppl. Table 13:** Distance-based redundancy analysis (db-RDA) for buccal samples. CP – chronic pancreatitis

| Variables | IRR | 95% CI | p-value |
|---|---|---|---|
| **Chronic pancreatitis** | | | |
| *No* | — | — | |
| *Yes* | 1.04 | 0.95, 1.14 | 0.4 |
| **Proton-pump inhibitors** | | | |
| *No* | — | — | |
| *Yes* | 1.01 | 0.97, 1.06 | 0.5 |
| **>10 cigarettes / day** | 1.02 | 0.96, 1.07 | 0.6 |
| **Etiology - Alcohol** | 0.96 | 0.91, 1.02 | 0.2 |

Abbreviations: CI = Confidence Interval, IRR = Incidence Rate Ratio

**Suppl. Table 14:** Multiple negative binomial regression for observed species in rectal samples. CI – Confidence interval, IRR – incidence rate ratio

| Variables | IRR | 95% CI | p-value |
|---|---|---|---|
| **Chronic pancreatitis** | | | |
| *No* | — | — | |
| *Yes* | 1.02 | 0.94, 1.12 | 0.6 |
| **Proton-pump inhibitors** | | | |
| *No* | — | — | |
| *Yes* | 1.04 | 1.00, 1.08 | 0.073 |
| **>10 cigarettes / day** | 1.01 | 0.96, 1.07 | 0.7 |
| **Etiology - Alcohol** | 0.96 | 0.92, 1.01 | 0.15 |

Abbreviation: CI = Confidence Interval

**Suppl. Table 15:** Multiple gamma regression for Shannon index in rectal samples. CI – Confidence interval, IRR – incidence rate ratio

| Variables | p-value |
|---|---|
| CP | 0.593 |
| Etio. - Alcohol | 0.046 |
| Proton-pump inhibitors | 0.405 |
| > 10 cigarettes / day | 0.018 |

**Suppl. Table 16:** Distance-based redundancy analysis (db-RDA) for rectal samples. CP – chronic pancreatitis

| Causes of post-discharge mortality | N = 277[1] |
|---|---|
| Total death count | 20 (7.2%) |
| Cancer related | 6 (2.2%) |
| Cardiovascular disease | 3 (1.1%) |
| Pancreatitis related | 1 (0.4%) |
| Sepsis or infection (not pancreatitis related) | 3 (1.1%) |
| Respiratory disease | 3 (1.1%) |
| Other | 2 (0.7%) |
| Not available | 3 (1.1%) |

[1]n (%)

**Suppl. Table 17:** Causes of post-discharge mortality

| **Post-discharge mortality** | **No** N = 220[1] | **Yes** N = 18[1] | **p-value**[2] |
|---|---|---|---|
| **Gender** | | | 0.6 |
| Female | 99 / 220 (45%) | 7 / 18 (39%) | |
| Male | 121 / 220 (55%) | 11 / 18 (61%) | |
| **Ethnicity** | | | >0.9 |
| African | 1 / 220 (0.5%) | 0 / 18 (0%) | |
| Asian | 1 / 220 (0.5%) | 0 / 18 (0%) | |
| Caucasian | 215 / 220 (98%) | 18 / 18 (100%) | |
| Hispanic or Latino | 3 / 220 (1.4%) | 0 / 18 (0%) | |
| **Etiology** | | | 0.7 |
| Alcoholic | 58 / 220 (26%) | 3 / 18 (17%) | |
| Biliary | 102 / 220 (46%) | 9 / 18 (50%) | |
| Idiopathic | 33 / 220 (15%) | 4 / 18 (22%) | |
| Other | 27 / 220 (12%) | 2 / 18 (11%) | |
| **Alcohol** | | | 0.4 |
| Current chronic alcohol consumption | 64 / 220 (29%) | 3 / 18 (17%) | |
| Former chronic alcohol consumption | 25 / 220 (11%) | 1 / 18 (5.6%) | |
| No alcohol consumption | 131 / 220 (60%) | 14 / 18 (78%) | |
| **Nicotine** | | | 0.3 |
| >10 cig./d | 48 / 220 (22%) | 2 / 18 (11%) | |
| 1-10 cig./d | 18 / 220 (8.2%) | 0 / 18 (0%) | |
| Non-smoker | 154 / 220 (70%) | 16 / 18 (89%) | |
| **Diet** | | | >0.9 |
| Omnivorous | 215 / 220 (98%) | 18 / 18 (100%) | |
| Vegetarian | 5 / 220 (2.3%) | 0 / 18 (0%) | |
| **Antibiotics** | | | **0.029** |
| Currently | 23 / 220 (10%) | 5 / 18 (28%) | |
| In the last seven days | 3 / 220 (1.4%) | 1 / 18 (5.6%) | |
| In the last six months | 17 / 220 (7.7%) | 2 / 18 (11%) | |
| More than six months ago | 177 / 220 (80%) | 10 / 18 (56%) | |
| **Cholestasis at index** | 51 / 220 (23%) | 5 / 18 (28%) | 0.8 |
| **No prior diseases** | 97 / 220 (44%) | 2 / 18 (11%) | **0.006** |
| **Cardiovascular disease** | 56 / 220 (25%) | 8 / 18 (44%) | 0.10 |
| **Heart insufficiency** | 14 / 220 (6.4%) | 5 / 18 (28%) | **0.008** |
| **Diabetes mellitus** | 31 / 220 (14%) | 4 / 18 (22%) | 0.3 |
| **Inflammatory bowel disease** | 3 / 220 (1.4%) | 1 / 18 (5.6%) | 0.3 |
| **Irritable bowel disease** | 4 / 220 (1.8%) | 0 / 18 (0%) | >0.9 |
| **Clostridioides diff. within last 12 month** | | | |
| No | 220 / 220 (100%) | 18 / 18 (100%) | |
| **Chronic constipation** | 3 / 220 (1.4%) | 3 / 18 (17%) | **0.006** |
| **Chronic diarrhea** | 1 / 220 (0.5%) | 0 / 18 (0%) | >0.9 |
| **MASLD** | 9 / 220 (4.1%) | 1 / 18 (5.6%) | 0.6 |
| **Liver cirrhosis** | 4 / 220 (1.8%) | 0 / 18 (0%) | >0.9 |
| **Other gastrointestinal disease** | 6 / 220 (2.7%) | 0 / 18 (0%) | >0.9 |
| **GI cancer** | 4 / 220 (1.8%) | 2 / 18 (11%) | 0.068 |
| **Non-GI cancer** | 12 / 220 (5.5%) | 5 / 18 (28%) | **0.005** |

| **Post-discharge mortality** | **No** N = 220[1] | **Yes** N = 18[1] | **p-value**[2] |
|---|---|---|---|
| **Hematologic cancer** | 2 / 214 (0.9%) | 1 / 17 (5.9%) | 0.2 |
| Unknown | 6 | 1 | |
| **Parkinson** | 0 / 220 (0%) | 1 / 18 (5.6%) | 0.076 |
| **Dementia** | 1 / 220 (0.5%) | 2 / 18 (11%) | **0.016** |
| **Depression** | 8 / 220 (3.6%) | 2 / 18 (11%) | 0.2 |
| **Autism** | | | |
| No | 220 / 220 (100%) | 18 / 18 (100%) | |
| **Other liver disease** | 3 / 220 (1.4%) | 0 / 18 (0%) | >0.9 |
| **GI surgery** | 4 / 220 (1.8%) | 0 / 18 (0%) | >0.9 |
| **Abdominal surgery** | 32 / 220 (15%) | 6 / 18 (33%) | **0.048** |
| **HIV** | | | |
| No | 220 / 220 (100%) | 18 / 18 (100%) | |
| **Rheumatic disease** | 8 / 220 (3.6%) | 1 / 18 (5.6%) | 0.5 |
| **No medication** | 74 / 220 (34%) | 6 / 18 (33%) | >0.9 |
| **Metformin** | 18 / 220 (8.2%) | 2 / 18 (11%) | 0.7 |
| **Other diabetes medication** | 9 / 220 (4.1%) | 0 / 18 (0%) | >0.9 |
| **Insulin** | 6 / 220 (2.7%) | 1 / 18 (5.6%) | 0.4 |
| **Proton-pump inhibitors** | 82 / 220 (37%) | 9 / 18 (50%) | 0.3 |
| **Immunosuppressants** | 6 / 220 (2.7%) | 1 / 18 (5.6%) | 0.4 |
| **Antidepressants** | 14 / 220 (6.4%) | 3 / 18 (17%) | 0.13 |
| **Neuroleptics** | 5 / 220 (2.3%) | 2 / 18 (11%) | 0.091 |
| **Paracetamol** | 27 / 220 (12%) | 3 / 18 (17%) | 0.5 |
| **NSAID** | 46 / 220 (21%) | 3 / 18 (17%) | >0.9 |
| **Opiates** | 25 / 220 (11%) | 2 / 18 (11%) | >0.9 |
| **Laxatives** | 6 / 220 (2.7%) | 3 / 18 (17%) | **0.023** |
| **Statins** | 40 / 220 (18%) | 7 / 18 (39%) | 0.058 |
| **Age** | 57 (18) | 73 (14) | **<0.001** |
| **BMI** | 29.6 (15.8) | 27.0 (4.4) | 0.3 |
| Unknown | 7 | 0 | |
| **Admission creatinine (mg/dl)** | 0.99 (0.61) | 1.13 (0.46) | **0.031** |
| Unknown | 7 | 0 | |

[1]n / N (%); Mean (SD)

[2]Pearson's Chi-squared test; Fisher's exact test; Wilcoxon rank sum test

**Suppl. Table 18:** Confounder distribution in buccal samples from patients that died after discharge and survivors. Differences were calculated using Fisher´s exact test, Chi-squared test or Mann-Whitney-U test. BMI – body mass index, GI – gastrointestinal, HIV – human immunodeficiency virus, MASLD – metabolic dysfunction-associated steatotic liver disease, NSAID – nonsteroidal anti-inflammatory drug, SD – standard deviation

| **Post-discharge mortality** | **No** N = 229[1] | **Yes** N = 20[1] | **p-value**[2] |
|---|---|---|---|
| **Gender** | | | >0.9 |
| Female | 104 / 229 (45%) | 9 / 20 (45%) | |
| Male | 125 / 229 (55%) | 11 / 20 (55%) | |
| **Ethnicity** | | | >0.9 |
| African | 1 / 229 (0.4%) | 0 / 20 (0%) | |
| Asian | 1 / 229 (0.4%) | 0 / 20 (0%) | |
| Caucasian | 223 / 229 (97%) | 20 / 20 (100%) | |
| Hispanic or Latino | 4 / 229 (1.7%) | 0 / 20 (0%) | |
| **Etiology** | | | 0.6 |
| Alcoholic | 59 / 229 (26%) | 3 / 20 (15%) | |
| Biliary | 111 / 229 (48%) | 11 / 20 (55%) | |
| Idiopathic | 30 / 229 (13%) | 4 / 20 (20%) | |
| Other | 29 / 229 (13%) | 2 / 20 (10%) | |
| **Alcohol** | | | 0.3 |
| Current chronic alcohol consumption | 63 / 229 (28%) | 3 / 20 (15%) | |
| Former chronic alcohol consumption | 30 / 229 (13%) | 1 / 20 (5.0%) | |
| No alcohol consumption | 136 / 229 (59%) | 16 / 20 (80%) | |
| **Nicotine** | | | 0.3 |
| >10 cig./d | 45 / 229 (20%) | 2 / 20 (10%) | |
| 1-10 cig./d | 19 / 229 (8.3%) | 0 / 20 (0%) | |
| Non-smoker | 165 / 229 (72%) | 18 / 20 (90%) | |
| **Diet** | | | >0.9 |
| Omnivorous | 224 / 229 (98%) | 20 / 20 (100%) | |
| Vegetarian | 5 / 229 (2.2%) | 0 / 20 (0%) | |
| **Antibiotics** | | | 0.084 |
| Currently | 32 / 229 (14%) | 5 / 20 (25%) | |
| In the last seven days | 6 / 229 (2.6%) | 2 / 20 (10%) | |
| In the last six months | 21 / 229 (9.2%) | 2 / 20 (10%) | |
| More than six months ago | 170 / 229 (74%) | 11 / 20 (55%) | |
| **Cholestasis at index** | 56 / 229 (24%) | 7 / 20 (35%) | 0.3 |
| **No prior diseases** | 93 / 229 (41%) | 2 / 20 (10%) | **0.007** |
| **Cardiovascular disease** | 56 / 229 (24%) | 10 / 20 (50%) | **0.013** |
| **Heart insufficiency** | 14 / 229 (6.1%) | 7 / 20 (35%) | **<0.001** |
| **Diabetes mellitus** | 32 / 229 (14%) | 6 / 20 (30%) | 0.10 |
| **Inflammatory bowel disease** | 3 / 229 (1.3%) | 1 / 20 (5.0%) | 0.3 |
| **Irritable bowel disease** | 6 / 229 (2.6%) | 0 / 20 (0%) | >0.9 |
| **Clostridioides diff. within last 12 month** | | | |
| No | 229 / 229 (100%) | 20 / 20 (100%) | |
| **Chronic constipation** | 4 / 229 (1.7%) | 3 / 20 (15%) | **0.013** |
| **Chronic diarrhea** | 3 / 229 (1.3%) | 0 / 20 (0%) | >0.9 |
| **MASLD** | 8 / 229 (3.5%) | 1 / 20 (5.0%) | 0.5 |
| **Liver cirrhosis** | 5 / 229 (2.2%) | 0 / 20 (0%) | >0.9 |
| **Other gastrointestinal disease** | 7 / 229 (3.1%) | 0 / 20 (0%) | >0.9 |
| **GI malignancy** | 4 / 229 (1.7%) | 2 / 20 (10%) | 0.076 |
| **Non-GI malignancy** | 14 / 229 (6.1%) | 5 / 20 (25%) | **0.011** |

| Post-discharge mortality | **No** N = 229[1] | **Yes** N = 20[1] | p-value[2] |
|---|---|---|---|
| **Hematologic malignancy** | 2 / 227 (0.9%) | 1 / 19 (5.3%) | 0.2 |
| Unknown | 2 | 1 | |
| **Parkinson** | 1 / 229 (0.4%) | 2 / 20 (10%) | **0.018** |
| **Dementia** | 1 / 229 (0.4%) | 2 / 20 (10%) | **0.018** |
| **Depression** | 11 / 229 (4.8%) | 2 / 20 (10%) | 0.3 |
| **Autism** | | | |
| No | 229 / 229 (100%) | 20 / 20 (100%) | |
| **Other liver disease** | 4 / 229 (1.7%) | 0 / 20 (0%) | >0.9 |
| **GI surgery** | 5 / 229 (2.2%) | 0 / 20 (0%) | >0.9 |
| **Abdominal surgery** | 39 / 229 (17%) | 6 / 20 (30%) | 0.2 |
| **HIV** | | | |
| No | 229 / 229 (100%) | 20 / 20 (100%) | |
| **Rheumatic disease** | 8 / 229 (3.5%) | 1 / 20 (5.0%) | 0.5 |
| **No medication** | 74 / 229 (32%) | 6 / 20 (30%) | 0.8 |
| **Metformin** | 18 / 229 (7.9%) | 4 / 20 (20%) | 0.086 |
| **Other diabetes medication** | 8 / 229 (3.5%) | 0 / 20 (0%) | >0.9 |
| **Insulin** | 6 / 229 (2.6%) | 1 / 20 (5.0%) | 0.4 |
| **Proton-pump inhibitors** | 94 / 229 (41%) | 11 / 20 (55%) | 0.2 |
| **Immunosuppressants** | 7 / 229 (3.1%) | 1 / 20 (5.0%) | 0.5 |
| **Antidepressants** | 16 / 229 (7.0%) | 3 / 20 (15%) | 0.2 |
| **Neuroleptics** | 4 / 229 (1.7%) | 2 / 20 (10%) | 0.076 |
| **Paracetamol** | 30 / 229 (13%) | 3 / 20 (15%) | 0.7 |
| **NSAID** | 53 / 229 (23%) | 4 / 20 (20%) | >0.9 |
| **Opiates** | 31 / 229 (14%) | 2 / 20 (10%) | >0.9 |
| **Laxatives** | 6 / 229 (2.6%) | 3 / 20 (15%) | **0.027** |
| **Statins** | 45 / 229 (20%) | 8 / 20 (40%) | **0.045** |
| **Age** | 57 (18) | 74 (13) | **<0.001** |
| **BMI** | 29.3 (15.5) | 27.1 (4.3) | 0.4 |
| Unknown | 8 | 0 | |
| **Admission creatinine (mg/dl)** | 0.98 (0.61) | 1.15 (0.45) | **0.008** |
| Unknown | 7 | 0 | |

[1]n / N (%); Mean (SD)

[2]Pearson's Chi-squared test; Fisher's exact test; Wilcoxon rank sum test

**Suppl. Table 19:** Confounder distribution in rectal samples from patients that died after discharge and survivors. Differences were calculated using Fisher´s exact test, Chi-squared test or Mann-Whitney-U test. BMI – body mass index, GI – gastrointestinal, HIV – human immunodeficiency virus, MASLD – metabolic dysfunction-associated steatotic liver disease, NSAID – nonsteroidal anti-inflammatory drug, SD – standard deviation

| Variables | IRR | 95% CI | p-value |
|---|---|---|---|
| **Death** | | | |
| *No* | — | — | |
| *Yes* | 0.90 | 0.80, 1.02 | 0.076 |
| **Heart insufficiency** | | | |
| *No* | — | — | |
| *Yes* | 1.09 | 0.97, 1.23 | 0.14 |
| **Age** | 1.00 | 1.00, 1.00 | 0.2 |
| **Non-GI cancer** | | | |
| *No* | — | — | |
| *Yes* | 1.00 | 0.89, 1.13 | >0.9 |

Abbreviations: CI = Confidence Interval, IRR = Incidence Rate Ratio

**Suppl. Table 20:** Multiple negative binomial regression for observed species in buccal samples. CI – confidence interval, GI – gastrointestinal, IRR – incidence rate ratio

| Variables | IRR | 95% CI[1] | p-value |
|---|---|---|---|
| **Death** | | | |
| *No* | — | — | |
| *Yes* | 0.88 | -0.41, 0.15 | 0.4 |
| **Heart insufficiency** | | | |
| *No* | — | — | |
| *Yes* | 1.38 | 0.04, 0.60 | **0.026** |
| **Age** | 1.00 | 0.00, 0.00 | 0.8 |
| **Non-GI cancer** | | | |
| *No* | — | — | |
| *Yes* | 1.10 | -0.19, 0.38 | 0.5 |

[1]CI = Confidence Interval

**Suppl. Table 21:** Multiple linear regression for Shannon index in buccal samples. CI – confidence interval, GI – gastrointestinal, Beta – regression coefficient

| Variables | p-value |
|---|---|
| Death | **0.042*** |
| Age | **0.001***** |
| Non-GI cancer | 0.43 |
| Heart insufficiency | 0.095 |

**Suppl. Table 22:** Distance-based redundancy analysis (db-RDA) for post-discharge mortality in buccal samples. GI – gastrointestinal

| Variables | IRR | 95% CI | p-value |
|---|---|---|---|
| **Death** | | | |
| *No* | — | — | |
| *Yes* | 1.03 | 0.96, 1.12 | 0.4 |
| **Non-GI malignancy** | | | |
| *No* | — | — | |
| *Yes* | 0.99 | 0.91, 1.07 | 0.7 |
| **Heart insufficiency** | | | |
| *No* | — | — | |
| *Yes* | 1.02 | 0.95, 1.11 | 0.6 |
| **Age** | 1.00 | 1.00, 1.00 | 0.9 |
| **Antibiotics in the last seven days** | 0.90 | 0.80, 1.01 | 0.073 |

Abbreviations: CI = Confidence Interval, IRR = Incidence Rate Ratio

**Suppl. Table 23:** Multiple negative binomial regression for observed species in rectal samples. CI – confidence interval, GI – gastrointestinal, IRR – incidence rate ratio

| Variables | IRR | 95% CI | p-value |
|---|---|---|---|
| **Death** | | | |
| *No* | — | — | |
| *Yes* | 1.01 | 0.93, 1.09 | 0.8 |
| **Non-GI malignancy** | | | |
| *No* | — | — | |
| *Yes* | 0.99 | 0.92, 1.07 | 0.8 |
| **Heart insufficiency** | | | |
| *No* | — | — | |
| *Yes* | 1.04 | 0.97, 1.13 | 0.3 |
| **Age** | 1.00 | 1.00, 1.00 | 0.8 |
| **Antibiotics in the last seven days** | 0.94 | 0.84, 1.05 | 0.3 |

Abbreviation: CI = Confidence Interval

**Suppl. Table 24:** Multiple gamma regression for Shannon index in rectal samples. CI – confidence interval, GI – gastrointestinal, IRR – incidence rate ratio

| Variables | p-value |
|---|---|
| Death | 0.156 |
| Age | **0.001*** ** |
| Non-GI malignancy | 0.856 |
| Heart insufficiency | 0.259 |
| Antibiotics in the last seven days | 0.473 |

**Suppl. Table 25:** Distance-based redundancy analysis (db-RDA) for post-discharge mortality in rectal samples. GI – gastrointestinal

| **Pancreatic exocrine insufficiency** | **No** N = 196[1] | **Yes** N = 16[1] | **p-value**[2] |
|---|---|---|---|
| **Gender** | | | 0.090 |
| Female | 92 / 196 (47%) | 4 / 16 (25%) | |
| Male | 104 / 196 (53%) | 12 / 16 (75%) | |
| **Ethnicity** | | | 0.2 |
| African | 0 / 196 (0%) | 1 / 16 (6.3%) | |
| Asian | 1 / 196 (0.5%) | 0 / 16 (0%) | |
| Caucasian | 193 / 196 (98%) | 15 / 16 (94%) | |
| Hispanic or Latino | 2 / 196 (1.0%) | 0 / 16 (0%) | |
| **Etiology** | | | 0.6 |
| Alcoholic | 47 / 196 (24%) | 4 / 16 (25%) | |
| Biliary | 95 / 196 (48%) | 7 / 16 (44%) | |
| Idiopathic | 28 / 196 (14%) | 4 / 16 (25%) | |
| Other | 26 / 196 (13%) | 1 / 16 (6.3%) | |
| **Alcohol** | | | **0.019** |
| Current chronic alcohol consumption | 53 / 196 (27%) | 4 / 16 (25%) | |
| Former chronic alcohol consumption | 15 / 196 (7.7%) | 5 / 16 (31%) | |
| No alcohol consumption | 128 / 196 (65%) | 7 / 16 (44%) | |
| **Nicotine** | | | 0.3 |
| >10 cig./d | 38 / 196 (19%) | 5 / 16 (31%) | |
| 1-10 cig./d | 15 / 196 (7.7%) | 0 / 16 (0%) | |
| Non-smoker | 143 / 196 (73%) | 11 / 16 (69%) | |
| **Diet** | | | >0.9 |
| Omnivorous | 193 / 196 (98%) | 16 / 16 (100%) | |
| Vegetarian | 3 / 196 (1.5%) | 0 / 16 (0%) | |
| **Antibiotics** | | | >0.9 |
| Currently | 25 / 196 (13%) | 1 / 16 (6.3%) | |
| In the last seven days | 4 / 196 (2.0%) | 0 / 16 (0%) | |
| In the last six months | 16 / 196 (8.2%) | 1 / 16 (6.3%) | |
| More than six months ago | 151 / 196 (77%) | 14 / 16 (88%) | |
| **Cholestasis at index** | 51 / 196 (26%) | 2 / 16 (13%) | 0.4 |
| **No prior diseases** | 76 / 196 (39%) | 7 / 16 (44%) | 0.7 |
| **Cardiovascular disease** | 53 / 196 (27%) | 5 / 16 (31%) | 0.8 |
| **Heart insufficiency** | 16 / 196 (8.2%) | 2 / 16 (13%) | 0.6 |
| **Diabetes mellitus** | 31 / 196 (16%) | 2 / 16 (13%) | >0.9 |
| **Inflammatory bowel disease** | 4 / 196 (2.0%) | 0 / 16 (0%) | >0.9 |
| **Irritable bowel disease** | 4 / 196 (2.0%) | 0 / 16 (0%) | >0.9 |
| **Clostridioides diff. within last 12 month** | | | |
| No | 196 / 196 (100%) | 16 / 16 (100%) | |
| **Chronic constipation** | 6 / 196 (3.1%) | 0 / 16 (0%) | >0.9 |
| **Chronic diarrhea** | 1 / 196 (0.5%) | 0 / 16 (0%) | >0.9 |
| **MASLD** | 9 / 196 (4.6%) | 0 / 16 (0%) | >0.9 |
| **Liver cirrhosis** | 4 / 196 (2.0%) | 0 / 16 (0%) | >0.9 |
| **Other gastrointestinal disease** | 5 / 196 (2.6%) | 0 / 16 (0%) | >0.9 |
| **GI cancer** | 5 / 196 (2.6%) | 1 / 16 (6.3%) | 0.4 |
| **Non-GI cancer** | 13 / 196 (6.6%) | 3 / 16 (19%) | 0.11 |

| **Pancreatic exocrine insufficiency** | **No** N = 196[1] | **Yes** N = 16[1] | **p-value**[2] |
|---|---|---|---|
| **Hematologic cancer** | 3 / 190 (1.6%) | 0 / 15 (0%) | >0.9 |
| Unknown | 6 | 1 | |
| **Parkinson** | 1 / 196 (0.5%) | 0 / 16 (0%) | >0.9 |
| **Dementia** | 3 / 196 (1.5%) | 0 / 16 (0%) | >0.9 |
| **Depression** | 9 / 196 (4.6%) | 1 / 16 (6.3%) | 0.6 |
| **Autism** | | | |
| No | 196 / 196 (100%) | 16 / 16 (100%) | |
| **Other liver disease** | 3 / 196 (1.5%) | 0 / 16 (0%) | >0.9 |
| **GI surgery** | 4 / 196 (2.0%) | 0 / 16 (0%) | >0.9 |
| **Abdominal surgery** | 34 / 196 (17%) | 0 / 16 (0%) | 0.081 |
| **HIV** | | | |
| No | 196 / 196 (100%) | 16 / 16 (100%) | |
| **Rheumatic disease** | 8 / 196 (4.1%) | 0 / 16 (0%) | >0.9 |
| **No medication** | 64 / 196 (33%) | 9 / 16 (56%) | 0.056 |
| **Metformin** | 17 / 196 (8.7%) | 2 / 16 (13%) | 0.6 |
| **Other diabetes medication** | 9 / 196 (4.6%) | 0 / 16 (0%) | >0.9 |
| **Insulin** | 7 / 196 (3.6%) | 0 / 16 (0%) | >0.9 |
| **Proton-pump inhibitors** | 76 / 196 (39%) | 3 / 16 (19%) | 0.11 |
| **Immunosuppressants** | 7 / 196 (3.6%) | 0 / 16 (0%) | >0.9 |
| **Antidepressants** | 16 / 196 (8.2%) | 1 / 16 (6.3%) | >0.9 |
| **Neuroleptics** | 7 / 196 (3.6%) | 0 / 16 (0%) | >0.9 |
| **Paracetamol** | 29 / 196 (15%) | 0 / 16 (0%) | 0.14 |
| **NSAID** | 40 / 196 (20%) | 0 / 16 (0%) | **0.046** |
| **Opiates** | 24 / 196 (12%) | 0 / 16 (0%) | 0.2 |
| **Laxatives** | 8 / 196 (4.1%) | 0 / 16 (0%) | >0.9 |
| **Statins** | 39 / 196 (20%) | 5 / 16 (31%) | 0.3 |
| **Age** | 58 (18) | 58 (14) | >0.9 |
| **BMI** | 28.6 (5.9) | 27.4 (4.4) | 0.5 |
| Unknown | 7 | 0 | |
| **Admission creatinine (mg/dl)** | 0.97 (0.57) | 1.17 (0.58) | 0.078 |
| Unknown | 2 | 0 | |

[1]n / N (%); Mean (SD)

[2]Pearson's Chi-squared test; Fisher's exact test; Wilcoxon rank sum test

**Suppl. Table 26:** Confounder distribution in buccal samples from patients with and without a pancreatic exocrine insufficiency. Differences were calculated using Fisher´s exact test, Chi-squared test or Mann-Whitney-U test. BMI – body mass index, GI – gastrointestinal, HIV – human immunodeficiency virus, MASLD – metabolic dysfunction-associated steatotic liver disease, NSAID – nonsteroidal anti-inflammatory drug, SD – standard deviation

| **Pancreatic exocrine insufficiency** | **No** N = 206[1] | **Yes** N = 15[1] | **p-value**[2] |
|---|---|---|---|
| **Gender** | | | 0.3 |
| Female | 98 / 206 (48%) | 5 / 15 (33%) | |
| Male | 108 / 206 (52%) | 10 / 15 (67%) | |
| **Ethnicity** | | | 0.14 |
| African | 0 / 206 (0%) | 1 / 15 (6.7%) | |
| Asian | 1 / 206 (0.5%) | 0 / 15 (0%) | |
| Caucasian | 202 / 206 (98%) | 14 / 15 (93%) | |
| Hispanic or Latino | 3 / 206 (1.5%) | 0 / 15 (0%) | |
| **Etiology** | | | 0.5 |
| Alcoholic | 49 / 206 (24%) | 3 / 15 (20%) | |
| Biliary | 104 / 206 (50%) | 7 / 15 (47%) | |
| Idiopathic | 25 / 206 (12%) | 4 / 15 (27%) | |
| Other | 28 / 206 (14%) | 1 / 15 (6.7%) | |
| **Alcohol** | | | **0.037** |
| Current chronic alcohol consumption | 54 / 206 (26%) | 3 / 15 (20%) | |
| Former chronic alcohol consumption | 19 / 206 (9.2%) | 5 / 15 (33%) | |
| No alcohol consumption | 133 / 206 (65%) | 7 / 15 (47%) | |
| **Nicotine** | | | 0.4 |
| >10 cig./d | 35 / 206 (17%) | 4 / 15 (27%) | |
| 1-10 cig./d | 16 / 206 (7.8%) | 0 / 15 (0%) | |
| Non-smoker | 155 / 206 (75%) | 11 / 15 (73%) | |
| **Diet** | | | >0.9 |
| Omnivorous | 203 / 206 (99%) | 15 / 15 (100%) | |
| Vegetarian | 3 / 206 (1.5%) | 0 / 15 (0%) | |
| **Antibiotics** | | | >0.9 |
| Currently | 33 / 206 (16%) | 2 / 15 (13%) | |
| In the last seven days | 8 / 206 (3.9%) | 0 / 15 (0%) | |
| In the last six months | 20 / 206 (9.7%) | 1 / 15 (6.7%) | |
| More than six months ago | 145 / 206 (70%) | 12 / 15 (80%) | |
| **Cholestasis at index** | 56 / 206 (27%) | 3 / 15 (20%) | 0.8 |
| **No prior diseases** | 71 / 206 (34%) | 8 / 15 (53%) | 0.14 |
| **Cardiovascular disease** | 55 / 206 (27%) | 4 / 15 (27%) | >0.9 |
| **Heart insufficiency** | 18 / 206 (8.7%) | 1 / 15 (6.7%) | >0.9 |
| **Diabetes mellitus** | 33 / 206 (16%) | 2 / 15 (13%) | >0.9 |
| **Inflammatory bowel disease** | 4 / 206 (1.9%) | 0 / 15 (0%) | >0.9 |
| **Irritable bowel disease** | 5 / 206 (2.4%) | 0 / 15 (0%) | >0.9 |
| **Clostridioides diff. within last 12 month** | | | |
| No | 206 / 206 (100%) | 15 / 15 (100%) | |
| **Chronic constipation** | 7 / 206 (3.4%) | 0 / 15 (0%) | >0.9 |
| **Chronic diarrhea** | 2 / 206 (1.0%) | 0 / 15 (0%) | >0.9 |
| **MASLD** | 9 / 206 (4.4%) | 0 / 15 (0%) | >0.9 |
| **Liver cirrhosis** | 5 / 206 (2.4%) | 0 / 15 (0%) | >0.9 |
| **Other gastrointestinal disease** | 6 / 206 (2.9%) | 0 / 15 (0%) | >0.9 |
| **GI malignancy** | 5 / 206 (2.4%) | 1 / 15 (6.7%) | 0.3 |
| **Non-GI malignancy** | 15 / 206 (7.3%) | 3 / 15 (20%) | 0.11 |

| Pancreatic exocrine insufficiency | **No** N = 206[1] | **Yes** N = 15[1] | p-value[2] |
|---|---|---|---|
| **Hematologic malignancy** | 3 / 203 (1.5%) | 0 / 15 (0%) | >0.9 |
| Unknown | 3 | 0 | |
| **Parkinson** | 3 / 206 (1.5%) | 0 / 15 (0%) | >0.9 |
| **Dementia** | 3 / 206 (1.5%) | 0 / 15 (0%) | >0.9 |
| **Depression** | 13 / 206 (6.3%) | 0 / 15 (0%) | 0.6 |
| **Autism** | | | |
| No | 206 / 206 (100%) | 15 / 15 (100%) | |
| **Other liver disease** | 4 / 206 (1.9%) | 0 / 15 (0%) | >0.9 |
| **GI surgery** | 5 / 206 (2.4%) | 0 / 15 (0%) | >0.9 |
| **Abdominal surgery** | 41 / 206 (20%) | 0 / 15 (0%) | 0.080 |
| **HIV** | | | |
| No | 206 / 206 (100%) | 15 / 15 (100%) | |
| **Rheumatic disease** | 9 / 206 (4.4%) | 0 / 15 (0%) | >0.9 |
| **No medication** | 63 / 206 (31%) | 10 / 15 (67%) | **0.008** |
| **Metformin** | 18 / 206 (8.7%) | 2 / 15 (13%) | 0.6 |
| **Other diabetes medication** | 8 / 206 (3.9%) | 0 / 15 (0%) | >0.9 |
| **Insulin** | 7 / 206 (3.4%) | 0 / 15 (0%) | >0.9 |
| **Proton-pump inhibitors** | 88 / 206 (43%) | 3 / 15 (20%) | 0.084 |
| **Immunosuppressants** | 8 / 206 (3.9%) | 0 / 15 (0%) | >0.9 |
| **Antidepressants** | 19 / 206 (9.2%) | 0 / 15 (0%) | 0.4 |
| **Neuroleptics** | 6 / 206 (2.9%) | 0 / 15 (0%) | >0.9 |
| **Paracetamol** | 32 / 206 (16%) | 0 / 15 (0%) | 0.14 |
| **NSAID** | 47 / 206 (23%) | 0 / 15 (0%) | **0.045** |
| **Opiates** | 29 / 206 (14%) | 0 / 15 (0%) | 0.2 |
| **Laxatives** | 8 / 206 (3.9%) | 0 / 15 (0%) | >0.9 |
| **Statins** | 44 / 206 (21%) | 4 / 15 (27%) | 0.7 |
| **Age** | 59 (18) | 58 (15) | 0.9 |
| **BMI** | 28.4 (5.5) | 27.2 (4.8) | 0.5 |
| Unknown | 8 | 0 | |
| **Admission creatinine (mg/dl)** | 0.96 (0.57) | 1.13 (0.62) | 0.3 |
| Unknown | 2 | 0 | |

[1]n / N (%); Mean (SD)

[2]Pearson's Chi-squared test; Fisher's exact test; Wilcoxon rank sum test

**Suppl. Table 27:** Confounder distribution in rectal samples from patients with and without a pancreatic exocrine insufficiency. Differences were calculated using Fisher´s exact test, Chi-squared test or Mann-Whitney-U test. BMI – body mass index, GI – gastrointestinal, HIV – human immunodeficiency virus, MASLD – metabolic dysfunction-associated steatotic liver disease, NSAID – nonsteroidal anti-inflammatory drug, SD – standard deviation

| Variables | IRR | 95% CI | p-value |
|---|---|---|---|
| **PEI** | | | |
| *No* | — | — | |
| *Yes* | 0.97 | 0.87, 1.10 | 0.6 |

Abbreviations: CI = Confidence Interval, IRR = Incidence Rate Ratio

**Suppl. Table 28:** Univariate analysis for observed species in buccal samples. CI – confidence interval, IRR – incidence rate ratio, PEI – pancreatic exocrine insufficiency

| Variables | IRR | 95% CI[1] | p-value |
|---|---|---|---|
| **PEI** | | | |
| *No* | — | — | |
| *Yes* | 0.92 | -0.36, 0.19 | 0.5 |

[1]CI = Confidence Interval

**Suppl. Table 29:** Univariate analysis for Shannon index in buccal samples.
CI – confidence interval, Beta – regression coefficient, PEI – pancreatic exocrine insufficiency

| Variables | p-value |
|---|---|
| PEI | 0.973 |

**Suppl. Table 30:** Bray-Curtis dissimilarity for pancreatic exocrine insufficiency (PEI) in buccal samples.

| Variables | IRR | 95% CI | p-value |
|---|---|---|---|
| **PEI** | | | |
| *No* | — | — | |
| *Yes* | 1.03 | 0.95, 1.13 | 0.4 |

Abbreviations: CI = Confidence Interval, IRR = Incidence Rate Ratio

**Suppl. Table 31:** Univariate analysis for observed species in rectal samples. CI – confidence interval, GI – gastrointestinal, IRR – incidence rate ratio, PEI – pancreatic exocrine insufficiency

| Variables | IRR | 95% CI | p-value |
|---|---|---|---|
| **PEI** | | | |
| *No* | — | — | |
| *Yes* | 1.03 | 0.95, 1.11 | 0.5 |

Abbreviation: CI = Confidence Interval

**Suppl. Table 32:** Univariate analysis for Shannon index in rectal samples. CI – confidence interval, GI – gastrointestinal, IRR – incidence rate ratio, PEI – pancreatic exocrine insufficiency

| Variables | p-value |
|---|---|
| PEI | 0.592 |

**Suppl. Table 33:** Bray-Curtis dissimilarity for pancreatic exocrine insufficiency (PEI) in rectal samples.

| **Developed diabetes** | **No** N = 179[1] | **Yes** N = 12[1] | **p-value**[2] |
|---|---|---|---|
| **Gender** | | | 0.8 |
| Female | 82 / 179 (46%) | 5 / 12 (42%) | |
| Male | 97 / 179 (54%) | 7 / 12 (58%) | |
| **Ethnicity** | | | >0.9 |
| African | 1 / 179 (0.6%) | 0 / 12 (0%) | |
| Asian | 1 / 179 (0.6%) | 0 / 12 (0%) | |
| Caucasian | 175 / 179 (98%) | 12 / 12 (100%) | |
| Hispanic or Latino | 2 / 179 (1.1%) | 0 / 12 (0%) | |
| **Etiology** | | | 0.8 |
| Alcoholic | 45 / 179 (25%) | 4 / 12 (33%) | |
| Biliary | 91 / 179 (51%) | 5 / 12 (42%) | |
| Idiopathic | 21 / 179 (12%) | 2 / 12 (17%) | |
| Other | 22 / 179 (12%) | 1 / 12 (8.3%) | |
| **Alcohol** | | | 0.3 |
| Current chronic alcohol consumption | 53 / 179 (30%) | 2 / 12 (17%) | |
| Former chronic alcohol consumption | 13 / 179 (7.3%) | 2 / 12 (17%) | |
| No alcohol consumption | 113 / 179 (63%) | 8 / 12 (67%) | |
| **Nicotine** | | | >0.9 |
| >10 cig./d | 39 / 179 (22%) | 3 / 12 (25%) | |
| 1-10 cig./d | 12 / 179 (6.7%) | 0 / 12 (0%) | |
| Non-smoker | 128 / 179 (72%) | 9 / 12 (75%) | |
| **Diet** | | | >0.9 |
| Omnivorous | 175 / 179 (98%) | 12 / 12 (100%) | |
| Vegetarian | 4 / 179 (2.2%) | 0 / 12 (0%) | |
| **Antibiotics** | | | 0.7 |
| Currently | 19 / 179 (11%) | 2 / 12 (17%) | |
| More than six months ago | 141 / 179 (79%) | 10 / 12 (83%) | |
| Within the last six months | 15 / 179 (8.4%) | 0 / 12 (0%) | |
| Within the last week | 4 / 179 (2.2%) | 0 / 12 (0%) | |
| **Cholestasis at index** | 45 / 179 (25%) | 4 / 12 (33%) | 0.5 |
| **No prior diseases** | 87 / 179 (49%) | 4 / 12 (33%) | 0.3 |
| **Cardiovascular disease** | 39 / 179 (22%) | 5 / 12 (42%) | 0.2 |
| **Heart insufficiency** | 7 / 179 (3.9%) | 2 / 12 (17%) | 0.10 |
| **Inflammatory bowel disease** | 3 / 179 (1.7%) | 0 / 12 (0%) | >0.9 |
| **Irritable bowel disease** | 3 / 179 (1.7%) | 0 / 12 (0%) | >0.9 |
| **Clostridioides diff. within last 12 month** | | | |
| No | 179 / 179 (100%) | 12 / 12 (100%) | |
| **Chronic constipation** | 4 / 179 (2.2%) | 0 / 12 (0%) | >0.9 |
| **Chronic diarrhea** | 1 / 179 (0.6%) | 0 / 12 (0%) | >0.9 |
| **MASLD** | 7 / 179 (3.9%) | 0 / 12 (0%) | >0.9 |
| **Liver cirrhosis** | 4 / 179 (2.2%) | 0 / 12 (0%) | >0.9 |
| **Other gastrointestinal disease** | 5 / 179 (2.8%) | 1 / 12 (8.3%) | 0.3 |
| **GI cancer** | 5 / 179 (2.8%) | 0 / 12 (0%) | >0.9 |
| **Non-GI cancer** | 12 / 179 (6.7%) | 1 / 12 (8.3%) | 0.6 |
| **Hematologic cancer** | 2 / 173 (1.2%) | 0 / 11 (0%) | >0.9 |

| **Developed diabetes** | **No** N = 179[1] | **Yes** N = 12[1] | **p-value**[2] |
|---|---|---|---|
| Unknown | 6 | 1 | |
| **Parkinson** | 1 / 179 (0.6%) | 0 / 12 (0%) | >0.9 |
| **Dementia** | 1 / 179 (0.6%) | 0 / 12 (0%) | >0.9 |
| **Depression** | 8 / 179 (4.5%) | 1 / 12 (8.3%) | 0.4 |
| **Autism** | | | |
| No | 179 / 179 (100%) | 12 / 12 (100%) | |
| **Other liver disease** | 3 / 179 (1.7%) | 0 / 12 (0%) | >0.9 |
| **GI surgery** | 4 / 179 (2.2%) | 0 / 12 (0%) | >0.9 |
| **Abdominal surgery** | 23 / 179 (13%) | 4 / 12 (33%) | 0.071 |
| **HIV** | | | |
| No | 179 / 179 (100%) | 12 / 12 (100%) | |
| **Rheumatic disease** | 8 / 179 (4.5%) | 0 / 12 (0%) | >0.9 |
| **No medication** | 73 / 179 (41%) | 4 / 12 (33%) | 0.8 |
| **Metformin** | | | |
| No | 179 / 179 (100%) | 12 / 12 (100%) | |
| **Other diabetes medication** | 1 / 179 (0.6%) | 0 / 12 (0%) | >0.9 |
| **Insulin** | | | |
| No | 179 / 179 (100%) | 12 / 12 (100%) | |
| **Proton-pump inhibitors** | 62 / 179 (35%) | 3 / 12 (25%) | 0.8 |
| **Immunosuppressants** | 3 / 179 (1.7%) | 1 / 12 (8.3%) | 0.2 |
| **Antidepressants** | 13 / 179 (7.3%) | 1 / 12 (8.3%) | >0.9 |
| **Neuroleptics** | 5 / 179 (2.8%) | 0 / 12 (0%) | >0.9 |
| **Paracetamol** | 24 / 179 (13%) | 0 / 12 (0%) | 0.4 |
| **NSAID** | 35 / 179 (20%) | 2 / 12 (17%) | >0.9 |
| **Opiates** | 22 / 179 (12%) | 0 / 12 (0%) | 0.4 |
| **Laxatives** | 8 / 179 (4.5%) | 0 / 12 (0%) | >0.9 |
| **Statins** | 23 / 179 (13%) | 4 / 12 (33%) | 0.071 |
| **Age** | 56 (18) | 58 (21) | 0.8 |
| **BMI** | 28.5 (5.7) | 26.5 (3.2) | 0.3 |
| Unknown | 4 | 0 | |
| **Admission creatinine (mg/dl)** | 0.95 (0.53) | 0.96 (0.38) | 0.5 |
| Unknown | 3 | 1 | |

[1]n / N (%); Mean (SD)

[2]Pearson's Chi-squared test; Fisher's exact test; Wilcoxon rank sum test

**Suppl. Table 34:** Confounder distribution in buccal samples from patients with and without a post-discharge diabetes mellitus. Differences were calculated using Fisher´s exact test, Chi-squared test or Mann-Whitney-U test. BMI – body mass index, GI – gastrointestinal, HIV – human immunodeficiency virus, MASLD – metabolic dysfunction-associated steatotic liver disease, NSAID – nonsteroidal anti-inflammatory drug, SD – standard deviation

| **Developed diabetes** | **No** N = 186[1] | **Yes** N = 12[1] | **p-value**[2] |
|---|---|---|---|
| **Gender** | | | 0.7 |
| Female | 87 / 186 (47%) | 5 / 12 (42%) | |
| Male | 99 / 186 (53%) | 7 / 12 (58%) | |
| **Ethnicity** | | | >0.9 |
| African | 1 / 186 (0.5%) | 0 / 12 (0%) | |
| Asian | 1 / 186 (0.5%) | 0 / 12 (0%) | |
| Caucasian | 181 / 186 (97%) | 12 / 12 (100%) | |
| Hispanic or Latino | 3 / 186 (1.6%) | 0 / 12 (0%) | |
| **Etiology** | | | >0.9 |
| Alcoholic | 48 / 186 (26%) | 3 / 12 (25%) | |
| Biliary | 96 / 186 (52%) | 6 / 12 (50%) | |
| Idiopathic | 19 / 186 (10%) | 2 / 12 (17%) | |
| Other | 23 / 186 (12%) | 1 / 12 (8.3%) | |
| **Alcohol** | | | 0.11 |
| Current chronic alcohol consumption | 52 / 186 (28%) | 2 / 12 (17%) | |
| Former chronic alcohol consumption | 15 / 186 (8.1%) | 3 / 12 (25%) | |
| No alcohol consumption | 119 / 186 (64%) | 7 / 12 (58%) | |
| **Nicotine** | | | >0.9 |
| >10 cig./d | 38 / 186 (20%) | 2 / 12 (17%) | |
| 1-10 cig./d | 13 / 186 (7.0%) | 0 / 12 (0%) | |
| Non-smoker | 135 / 186 (73%) | 10 / 12 (83%) | |
| **Diet** | | | >0.9 |
| Omnivorous | 182 / 186 (98%) | 12 / 12 (100%) | |
| Vegetarian | 4 / 186 (2.2%) | 0 / 12 (0%) | |
| **Antibiotics** | | | 0.8 |
| Currently | 25 / 186 (13%) | 2 / 12 (17%) | |
| In the last seven days | 6 / 186 (3.2%) | 0 / 12 (0%) | |
| In the last six months | 19 / 186 (10%) | 0 / 12 (0%) | |
| More than six months ago | 136 / 186 (73%) | 10 / 12 (83%) | |
| **Cholestasis at index** | 47 / 186 (25%) | 5 / 12 (42%) | 0.3 |
| **No prior diseases** | 83 / 186 (45%) | 4 / 12 (33%) | 0.4 |
| **Cardiovascular disease** | 37 / 186 (20%) | 6 / 12 (50%) | **0.025** |
| **Heart insufficiency** | 6 / 186 (3.2%) | 1 / 12 (8.3%) | 0.4 |
| **Inflammatory bowel disease** | 3 / 186 (1.6%) | 0 / 12 (0%) | >0.9 |
| **Irritable bowel disease** | 5 / 186 (2.7%) | 0 / 12 (0%) | >0.9 |
| **Clostridioides diff. within last 12 month** | | | |
| No | 186 / 186 (100%) | 12 / 12 (100%) | |
| **Chronic constipation** | 5 / 186 (2.7%) | 0 / 12 (0%) | >0.9 |
| **Chronic diarrhea** | 3 / 186 (1.6%) | 0 / 12 (0%) | >0.9 |
| **MASLD** | 5 / 186 (2.7%) | 0 / 12 (0%) | >0.9 |
| **Liver cirrhosis** | 5 / 186 (2.7%) | 0 / 12 (0%) | >0.9 |
| **Other gastrointestinal disease** | 6 / 186 (3.2%) | 1 / 12 (8.3%) | 0.4 |
| **GI malignancy** | 5 / 186 (2.7%) | 0 / 12 (0%) | >0.9 |
| **Non-GI malignancy** | 13 / 186 (7.0%) | 2 / 12 (17%) | 0.2 |
| **Hematologic malignancy** | 2 / 183 (1.1%) | 0 / 12 (0%) | >0.9 |

| Developed diabetes | No N = 186[1] | Yes N = 12[1] | p-value[2] |
|---|---|---|---|
| Unknown | 3 | 0 | |
| **Parkinson** | 2 / 186 (1.1%) | 0 / 12 (0%) | >0.9 |
| **Dementia** | 1 / 186 (0.5%) | 0 / 12 (0%) | >0.9 |
| **Depression** | 11 / 186 (5.9%) | 1 / 12 (8.3%) | 0.5 |
| **Autism** | | | |
| No | 186 / 186 (100%) | 12 / 12 (100%) | |
| **Other liver disease** | 4 / 186 (2.2%) | 0 / 12 (0%) | >0.9 |
| **GI surgery** | 4 / 186 (2.2%) | 0 / 12 (0%) | >0.9 |
| **Abdominal surgery** | 29 / 186 (16%) | 4 / 12 (33%) | 0.12 |
| **HIV** | | | |
| No | 186 / 186 (100%) | 12 / 12 (100%) | |
| **Rheumatic disease** | 7 / 186 (3.8%) | 0 / 12 (0%) | >0.9 |
| **No medication** | 73 / 186 (39%) | 4 / 12 (33%) | 0.8 |
| **Metformin** | | | |
| No | 186 / 186 (100%) | 12 / 12 (100%) | |
| **Other diabetes medication** | | | |
| No | 186 / 186 (100%) | 12 / 12 (100%) | |
| **Insulin** | | | |
| No | 186 / 186 (100%) | 12 / 12 (100%) | |
| **Proton-pump inhibitors** | 70 / 186 (38%) | 5 / 12 (42%) | 0.8 |
| **Immunosuppressants** | 4 / 186 (2.2%) | 1 / 12 (8.3%) | 0.3 |
| **Antidepressants** | 15 / 186 (8.1%) | 1 / 12 (8.3%) | >0.9 |
| **Neuroleptics** | 4 / 186 (2.2%) | 0 / 12 (0%) | >0.9 |
| **Paracetamol** | 26 / 186 (14%) | 0 / 12 (0%) | 0.4 |
| **NSAID** | 39 / 186 (21%) | 2 / 12 (17%) | >0.9 |
| **Opiates** | 26 / 186 (14%) | 1 / 12 (8.3%) | >0.9 |
| **Laxatives** | 8 / 186 (4.3%) | 0 / 12 (0%) | >0.9 |
| **Statins** | 24 / 186 (13%) | 4 / 12 (33%) | 0.071 |
| **Age** | 57 (18) | 59 (21) | 0.7 |
| **BMI** | 28.2 (5.5) | 26.4 (3.2) | 0.3 |
| Unknown | 5 | 0 | |
| **Admission creatinine (mg/dl)** | 0.96 (0.56) | 0.90 (0.37) | 0.8 |
| Unknown | 3 | 1 | |

[1]n / N (%); Mean (SD)

[2]Pearson's Chi-squared test; Fisher's exact test; Wilcoxon rank sum test

**Suppl. Table 35:** Confounder distribution in rectal samples from patients with and without a post-discharge diabetes mellitus. Differences were calculated using Fisher´s exact test, Chi-squared test or Mann-Whitney-U test. BMI – body mass index, GI – gastrointestinal, HIV – human immunodeficiency virus, MASLD – metabolic dysfunction-associated steatotic liver disease, NSAID – nonsteroidal anti-inflammatory drug, SD – standard deviation

| Variables | IRR | 95% CI | p-value |
|---|---|---|---|
| **Diabetes** | | | |
| *No* | — | — | |
| *Yes* | 0.93 | 0.82, 1.07 | 0.3 |

Abbreviations: CI = Confidence Interval, IRR = Incidence Rate Ratio

**Suppl. Table 36:** Univariate analysis for observed species in buccal samples. CI – confidence interval, IRR – incidence rate ratio

| Variables | IRR | 95% CI[1] | p-value |
|---|---|---|---|
| **Diabetes** | | | |
| *No* | — | — | |
| *Yes* | 0.78 | -0.56, 0.07 | 0.13 |

[1]CI = Confidence Interval

**Suppl. Table 37:** Univariate analysis for Shannon index in buccal samples. CI – confidence interval, Beta – regression coefficient

| Variables | p-value |
|---|---|
| Post discharge diabetes mellitus | 0.082 |

**Suppl. Table 38:** Bray-Curtis dissimilarity for post discharge diabetes mellitus in buccal samples.

| Variables | IRR | 95% CI | p-value |
|---|---|---|---|
| **Diabetes** | | | |
| *No* | — | — | |
| *Yes* | 0.97 | 0.88, 1.08 | 0.6 |
| **Cardiovascular disease** | | | |
| *No* | — | — | |
| *Yes* | 0.98 | 0.93, 1.04 | 0.5 |

Abbreviations: CI = Confidence Interval, IRR = Incidence Rate Ratio

**Suppl. Table 39:** Multiple negative binomial regression for observed species in rectal samples. CI – confidence interval, IRR – incidence rate ratio

| Variables | IRR | 95% CI | p-value |
|---|---|---|---|
| **Diabetes** | | | |
| *No* | — | — | |
| *Yes* | 0.93 | 0.84, 1.02 | 0.11 |
| **Cardiovascular disease** | | | |
| *No* | — | — | |
| *Yes* | 1.00 | 0.95, 1.06 | >0.9 |

Abbreviation: CI = Confidence Interval

**Suppl. Table 40:** Multiple gamma regression for Shannon index in rectal samples. CI – confidence interval, IRR – incidence rate ratio

| **Variables** | **p-value** |
|---|---|
| Post discharge diabetes mellitus | **0.026*** |
| Cardiovascular disease | 0.385 |

**Suppl. Table 41:** Bray-Curtis dissimilarity for post discharge diabetes mellitus in rectal samples.

| **Recurrent acute pancreatitis** | **0** N = 81[1] | **1** N = 53[1] | **p-value** |
|---|---|---|---|
| **No prior diseases** | 33 / 81 (41%) | 20 / 53 (38%) | 0.7 |
| **Cardiovascular disease** | 24 / 81 (30%) | 13 / 53 (25%) | 0.5 |
| **Heart insufficiency** | 5 / 81 (6.2%) | 5 / 53 (9.4%) | 0.5 |
| **Diabetes mellitus** | 13 / 81 (16%) | 10 / 53 (19%) | 0.7 |
| **Inflammatory bowel disease** | 1 / 81 (1.2%) | 0 / 53 (0%) | >0.9 |
| **Irritable bowel disease** | 0 / 81 (0%) | 2 / 53 (3.8%) | 0.2 |
| **Clostridioides diff. within last 12 month** | | | |
| No | 81 / 81 (100%) | 53 / 53 (100%) | |
| **Chronic constipation** | 4 / 81 (4.9%) | 2 / 53 (3.8%) | >0.9 |
| **Chronic diarrhea** | 1 / 81 (1.2%) | 1 / 53 (1.9%) | >0.9 |
| **MAFLD** | 3 / 81 (3.7%) | 1 / 53 (1.9%) | >0.9 |
| **Liver cirrhosis** | 2 / 81 (2.5%) | 2 / 53 (3.8%) | 0.6 |
| **Other gastrointestinal disease** | 3 / 81 (3.7%) | 3 / 53 (5.7%) | 0.7 |
| **GI cancer** | 1 / 81 (1.2%) | 1 / 53 (1.9%) | >0.9 |
| **Non-GI cancer** | 6 / 81 (7.4%) | 6 / 53 (11%) | 0.5 |
| **Hematologic cancer** | 2 / 80 (2.5%) | 0 / 52 (0%) | 0.5 |
| Unknown | 1 | 1 | |
| **Parkinson** | 2 / 81 (2.5%) | 0 / 53 (0%) | 0.5 |
| **Dementia** | 1 / 81 (1.2%) | 1 / 53 (1.9%) | >0.9 |
| **Depression** | 6 / 81 (7.4%) | 1 / 53 (1.9%) | 0.2 |
| **Autism** | | | |
| No | 81 / 81 (100%) | 53 / 53 (100%) | |
| **Other liver disease** | 2 / 81 (2.5%) | 0 / 53 (0%) | 0.5 |
| **GI surgery** | 1 / 81 (1.2%) | 1 / 53 (1.9%) | >0.9 |
| **Abdominal surgery** | 12 / 81 (15%) | 11 / 53 (21%) | 0.4 |
| **HIV** | | | |
| No | 81 / 81 (100%) | 53 / 53 (100%) | |
| **Rheumatic disease** | 3 / 81 (3.7%) | 0 / 53 (0%) | 0.3 |
| **No medication** | 23 / 81 (28%) | 19 / 53 (36%) | 0.4 |
| **Metformin** | 5 / 81 (6.2%) | 7 / 53 (13%) | 0.2 |
| **Other diabetes medication** | 1 / 81 (1.2%) | 4 / 53 (7.5%) | 0.080 |
| **Insulin** | 3 / 81 (3.7%) | 0 / 53 (0%) | 0.3 |
| **Proton-pump inhibitors** | 32 / 81 (40%) | 18 / 53 (34%) | 0.5 |
| **Immunosuppressant** | 4 / 81 (4.9%) | 0 / 53 (0%) | 0.2 |
| **Antidepressants** | 9 / 81 (11%) | 1 / 53 (1.9%) | 0.088 |
| **Neuroleptics** | 2 / 81 (2.5%) | 1 / 53 (1.9%) | >0.9 |
| **Paracetamol** | 8 / 81 (9.9%) | 6 / 53 (11%) | 0.8 |
| **NSAID** | 20 / 81 (25%) | 12 / 53 (23%) | 0.8 |
| **Opiates*** | 16 / 81 (20%) | 3 / 53 (5.7%) | **0.022** |
| **Laxatives** | 3 / 81 (3.7%) | 4 / 53 (7.5%) | 0.4 |
| **Statins** | 22 / 81 (27%) | 10 / 53 (19%) | 0.3 |
| **Ethnicity** | | | 0.8 |
| African | 1 / 81 (1.2%) | 0 / 53 (0%) | |
| Asian | 1 / 81 (1.2%) | 0 / 53 (0%) | |
| Caucasian | 77 / 81 (95%) | 53 / 53 (100%) | |

| **Recurrent acute pancreatitis** | **0** N = 81[1] | **1** N = 53[1] | **p-value** |
|---|---|---|---|
| Hispanic or Latino | 2 / 81 (2.5%) | 0 / 53 (0%) | |
| **Alcohol*** | | | 0.7 |
| Current chronic alcohol consumption | 31 / 81 (38%) | 24 / 53 (45%) | |
| Former chronic alcohol consumption | 12 / 81 (15%) | 8 / 53 (15%) | |
| No alcohol consumption | 38 / 81 (47%) | 21 / 53 (40%) | |
| **Gender** | | | 0.4 |
| Female | 23 / 81 (28%) | 19 / 53 (36%) | |
| Male | 58 / 81 (72%) | 34 / 53 (64%) | |
| **Diet** | | | 0.6 |
| Omnivorous | 80 / 81 (99%) | 51 / 53 (96%) | |
| Vegetarian | 1 / 81 (1.2%) | 2 / 53 (3.8%) | |
| **Antibiotics*** | | | >0.9 |
| >6month | 70 / 81 (86%) | 47 / 53 (89%) | |
| 6month | 5 / 81 (6.2%) | 3 / 53 (5.7%) | |
| current | 6 / 81 (7.4%) | 3 / 53 (5.7%) | |
| **Nicotine*** | | | 0.4 |
| >10 cig./d | 20 / 81 (25%) | 17 / 53 (32%) | |
| 1-10 cig./d | 6 / 81 (7.4%) | 6 / 53 (11%) | |
| No Smoker | 55 / 81 (68%) | 30 / 53 (57%) | |
| **Etiology** | | | 0.4 |
| Alcoholic | 31 / 81 (38%) | 23 / 53 (43%) | |
| Biliary | 29 / 81 (36%) | 16 / 53 (30%) | |
| Drug induced | 3 / 81 (3.7%) | 0 / 53 (0%) | |
| Hypertriglyceridemia | 0 / 81 (0%) | 2 / 53 (3.8%) | |
| Iatrogenic | 4 / 81 (4.9%) | 1 / 53 (1.9%) | |
| Idiopathic | 10 / 81 (12%) | 9 / 53 (17%) | |
| Other | 4 / 81 (4.9%) | 2 / 53 (3.8%) | |
| **Cholestasis at index*** | 12 / 81 (15%) | 6 / 53 (11%) | 0.6 |
| **Age** | 57 (17) | 55 (17) | 0.5 |
| **BMI** | 28.5 (5.9) | 27.6 (4.8) | 0.6 |
| Unknown | 2 | 2 | |
| **Admission Creatinine (mg/dl)** | 1.10 (0.83) | 1.02 (0.43) | 0.5 |
| Unknown | 2 | 3 | |

[1]n / N (%); Mean (SD)

**Suppl. Table 42:** Confounder distribution in matched rectal samples from patients with recurrent acute pancreatitis (RAP) and non-RAP. Differences were calculated using Fisher´s exact test, Chi-squared test or Mann-Whitney-U test. BMI – body mass index, GI – gastrointestinal, HIV – human immunodeficiency virus, MASLD – metabolic dysfunction-associated steatotic liver disease, NSAID – nonsteroidal anti-inflammatory drug, SD – standard deviation. *marked variables were used for matching.

| **Species** | **Cohort** | **Method** | **p value MaAsLin2** | **p value LEfSe** |
|---|---|---|---|---|
| *Butyricimonas faecihominis* | Non-RAP | MaAsLin2 & LEfSe | 0.001 | 0.001 |
| *Phocaeicola vulgatus* | Non-RAP | MaAsLin2 & LEfSe | 0.003 | 0.01 |

| | | | | |
|---|---|---|---|---|
| *Faecalibacterium prausnitzii* | Non-RAP | MaAsLin2 & LEfSe | 0.003 | 0.015 |
| *Coprococcus comes* | Non-RAP | MaAsLin2 & LEfSe | 0.004 | 0.013 |
| *Parabacteroides distasonis* | Non-RAP | MaAsLin2 & LEfSe | 0.008 | 0.049 |
| *Blautia obeum* | Non-RAP | MaAsLin2 & LEfSe | 0.013 | 0.03 |
| *Faecalibacterium duncaniae* | Non-RAP | MaAsLin2 & LEfSe | 0.012 | 0.022 |
| *Segatella copri* | Non-RAP | MaAsLin2 & LEfSe | 0.011 | 0.001 |
| *Blautia luti* | Non-RAP | MaAsLin2 & LEfSe | 0.034 | 0.039 |
| *Mobiluncus massiliensis* | Non-RAP | MaAsLin2 & LEfSe | 0.034 | 0.011 |
| *Anaerococcus prevotii* | RAP | MaAsLin2 | 0.016 | |
| *Anaerococcus mediterraneensis* | RAP | LefSe | | 0.037 |
| *Peptoniphilus harei* | RAP | LefSe | | 0.043 |
| *Finegoldia magna* | RAP | LefSe | | 0.028 |

**Suppl. Table 43:** Differential abundance between patients with and without recurrent acute pancreatitis (RAP) was calculated with Linear discriminant analysis effect size (LEfSe) and Microbiome Multivariable Association with Linear Models (MaAsLin2). Median abundance of one group was 0.1%.

| Intervention | **0** N = 33[1] | **1** N = 17[1] | p-value |
|---|---|---|---|
| **No prior diseases*** | 4 / 33 (12%) | 2 / 17 (12%) | >0.9 |
| **Cardiovascular disease** | 17 / 33 (52%) | 7 / 17 (41%) | 0.5 |
| **Heart insufficiency*** | 7 / 33 (21%) | 4 / 17 (24%) | >0.9 |
| **Diabetes mellitus** | 10 / 33 (30%) | 3 / 17 (18%) | 0.5 |
| **Inflammatory bowel disease** | 2 / 33 (6.1%) | 1 / 17 (5.9%) | >0.9 |
| **Irritable bowel disease** | 1 / 33 (3.0%) | 0 / 17 (0%) | >0.9 |
| **Clostridioides diff. within last 12 month** | | | |
| No | 33 / 33 (100%) | 17 / 17 (100%) | |
| **Chronic constipation** | 0 / 33 (0%) | 2 / 17 (12%) | 0.11 |
| **Chronic diarrhea** | | | |
| No | 33 / 33 (100%) | 17 / 17 (100%) | |
| **MAFLD** | 0 / 33 (0%) | 1 / 17 (5.9%) | 0.3 |
| **Liver cirrhosis** | | | |
| No | 33 / 33 (100%) | 17 / 17 (100%) | |
| **Other gastrointestinal disease** | 1 / 33 (3.0%) | 0 / 17 (0%) | >0.9 |
| **GI cancer** | 1 / 33 (3.0%) | 2 / 17 (12%) | 0.3 |
| **Non-GI solid cancer*** | 3 / 33 (9.1%) | 5 / 17 (29%) | 0.10 |
| **Hematologic cancer** | 1 / 33 (3.0%) | 1 / 16 (6.3%) | >0.9 |
| Unknown | 0 | 1 | |
| **Parkinson** | 0 / 33 (0%) | 1 / 17 (5.9%) | 0.3 |
| **Dementia** | 1 / 33 (3.0%) | 1 / 17 (5.9%) | >0.9 |
| **Depression** | 0 / 33 (0%) | 2 / 17 (12%) | 0.11 |
| **Autism** | | | |
| No | 33 / 33 (100%) | 17 / 17 (100%) | |
| **Other liver disease** | 2 / 33 (6.1%) | 0 / 17 (0%) | 0.5 |
| **GI surgery** | 1 / 33 (3.0%) | 0 / 17 (0%) | >0.9 |
| **Abdominal surgery** | 5 / 33 (15%) | 5 / 17 (29%) | 0.3 |
| **HIV** | | | |
| No | 33 / 33 (100%) | 17 / 17 (100%) | |
| **Rheumatic disease** | 2 / 33 (6.1%) | 0 / 17 (0%) | 0.5 |
| **No medication** | 9 / 33 (27%) | 6 / 17 (35%) | 0.6 |
| **Metformin** | 4 / 33 (12%) | 2 / 17 (12%) | >0.9 |
| **Other diabetes medication** | 4 / 33 (12%) | 0 / 17 (0%) | 0.3 |
| **Insulin** | 4 / 33 (12%) | 1 / 17 (5.9%) | 0.6 |
| **Proton-pump inhibitors** | 15 / 33 (45%) | 8 / 17 (47%) | >0.9 |
| **Immunosuppressant** | 1 / 33 (3.0%) | 1 / 17 (5.9%) | >0.9 |
| **Antidepressants** | 3 / 33 (9.1%) | 2 / 17 (12%) | >0.9 |
| **Neuroleptics** | 3 / 33 (9.1%) | 1 / 17 (5.9%) | >0.9 |
| **Paracetamol** | 5 / 33 (15%) | 3 / 17 (18%) | >0.9 |
| **NSAID** | 5 / 33 (15%) | 3 / 17 (18%) | >0.9 |
| **Opiates** | 1 / 33 (3.0%) | 1 / 17 (5.9%) | >0.9 |
| **Laxatives** | 0 / 33 (0%) | 2 / 17 (12%) | 0.11 |
| **Statins** | 10 / 33 (30%) | 6 / 17 (35%) | 0.7 |
| **Ethnicity** | | | >0.9 |

| Intervention | **0** N = 33[1] | **1** N = 17[1] | p-value |
|---|---|---|---|
| Asian | 1 / 33 (3.0%) | 0 / 17 (0%) | |
| Caucasian | 31 / 33 (94%) | 17 / 17 (100%) | |
| Hispanic or Latino | 1 / 33 (3.0%) | 0 / 17 (0%) | |
| **Alcohol** | | | 0.6 |
| Current chronic alcohol consumption | 9 / 33 (27%) | 3 / 17 (18%) | |
| Former chronic alcohol consumption | 4 / 33 (12%) | 1 / 17 (5.9%) | |
| No alcohol consumption | 20 / 33 (61%) | 13 / 17 (76%) | |
| **Gender** | | | 0.8 |
| Female | 13 / 33 (39%) | 6 / 17 (35%) | |
| Male | 20 / 33 (61%) | 11 / 17 (65%) | |
| **Diet** | | | |
| Omnivorous | 33 / 33 (100%) | 17 / 17 (100%) | |
| **Antibiotics** | | | >0.9 |
| >6month | 20 / 33 (61%) | 10 / 17 (59%) | |
| 6month | 1 / 33 (3.0%) | 1 / 17 (5.9%) | |
| current | 10 / 33 (30%) | 5 / 17 (29%) | |
| week | 2 / 33 (6.1%) | 1 / 17 (5.9%) | |
| **Nicotine** | | | >0.9 |
| >10 cig./d | 4 / 33 (12%) | 2 / 17 (12%) | |
| 1-10 cig./d | 1 / 33 (3.0%) | 0 / 17 (0%) | |
| No Smoker | 28 / 33 (85%) | 15 / 17 (88%) | |
| **Etiology** | | | 0.6 |
| Alcoholic | 5 / 33 (15%) | 3 / 17 (18%) | |
| Biliary | 20 / 33 (61%) | 8 / 17 (47%) | |
| Drug induced | 0 / 33 (0%) | 1 / 17 (5.9%) | |
| Iatrogenic | 3 / 33 (9.1%) | 1 / 17 (5.9%) | |
| Idiopathic | 5 / 33 (15%) | 4 / 17 (24%) | |
| **Cholestasis at index** | 10 / 33 (30%) | 4 / 17 (24%) | 0.7 |
| **Age*** | 67 (18) | 72 (14) | 0.4 |
| **BMI** | 28.7 (4.4) | 27.2 (4.5) | 0.3 |
| Unknown | 3 | 0 | |
| **Admission Creatinine (mg/dl)** | 1.15 (0.80) | 1.14 (0.47) | 0.5 |
| Unknown | 1 | 0 | |

[1]n / N (%); Mean (SD)

**Suppl. Table 44:** Confounder distribution in matched buccal samples from patients died after discharge and survivors. Differences were calculated using Fisher´s exact test, Chi-squared test or Mann-Whitney-U test. BMI – body mass index, GI – gastrointestinal, HIV – human immunodeficiency virus, MASLD – metabolic dysfunction-associated steatotic liver disease, NSAID – nonsteroidal anti-inflammatory drug, SD – standard deviation. *marked variables were used for matching.

| **Species** | **Cohort** | **Method** | **p value MaAsLin2** | **p value LEfSe** |
|---|---|---|---|---|
| *Rummeliibacillus stabekisii* | Dead | MaAsLin2 | 0.032 | |
| *Lactobacillus paragasseri* | Dead | LefSe | | 0.016 |

| | | | | |
|---|---|---|---|---|
| *Peptacetobacter hiranonis* | Survivors | MaAsLin2 | 0.017 | |
| *Streptococcus sp. FSL W8-0197* | Survivors | MaAsLin2 & LEfSe | 0.027 | 0.034 |
| *Klebsiella electrica* | Survivors | MaAsLin2 | 0.028 | |
| *Gemella morbillorum* | Survivors | MaAsLin2 & LEfSe | 0.029 | 0.028 |
| *Terrisporobacter petrolearius* | Survivors | MaAsLin2 | 0.031 | |
| *Clostridioides difficile* | Survivors | MaAsLin2 | 0.035 | |
| *Exiguobacterium alkaliphilum* | Survivors | MaAsLin2 | 0.043 | |
| *Paraclostridium sordellii* | Survivors | MaAsLin2 | 0.047 | |
| *Streptococcus gwangjuense* | Survivors | LefSe | | 0.042 |

**Suppl. Table 45:** Differential abundance between survivors and patients who died after discharge was calculated with Linear discriminant analysis effect size (LEfSe) and Microbiome Multivariable Association with Linear Models (MaAsLin2).

| **Post-discharge diabetes mellitus** | **No DM** N = 30[1] | **DM** N = 11[1] | **p-value** |
|---|---|---|---|
| **No prior diseases** | 18 / 30 (60%) | 7 / 11 (64%) | >0.9 |
| **Cardiovascular disease*** | 15 / 30 (50%) | 6 / 11 (55%) | 0.8 |
| **Heart insufficiency** | 30 / 30 (100%) | 10 / 11 (91%) | 0.3 |
| **Diabetes mellitus** | | | |
| Yes | 30 / 30 (100%) | 11 / 11 (100%) | |
| **Inflammatory bowel disease** | 29 / 30 (97%) | 11 / 11 (100%) | >0.9 |
| **Irritable bowel disease** | | | |
| Yes | 30 / 30 (100%) | 11 / 11 (100%) | |
| **Clostridioides diff. within last 12 month** | | | |
| Yes | 30 / 30 (100%) | 11 / 11 (100%) | |
| **Chronic constipation** | 29 / 30 (97%) | 11 / 11 (100%) | >0.9 |
| **Chronic diarrhea** | | | |
| Yes | 30 / 30 (100%) | 11 / 11 (100%) | |
| **MAFLD** | | | |
| Yes | 30 / 30 (100%) | 11 / 11 (100%) | |
| **Liver cirrhosis** | | | |
| Yes | 30 / 30 (100%) | 11 / 11 (100%) | |
| **Other gastrointestinal disease** | 29 / 30 (97%) | 10 / 11 (91%) | 0.5 |
| **GI cancer** | | | |
| Yes | 30 / 30 (100%) | 11 / 11 (100%) | |
| **Non-GI cancer** | 29 / 30 (97%) | 10 / 11 (91%) | 0.5 |
| **Hematologic cancer** | | | |
| Yes | 30 / 30 (100%) | 11 / 11 (100%) | |
| **Parkinson** | | | |
| Yes | 30 / 30 (100%) | 11 / 11 (100%) | |
| **Dementia** | | | |
| Yes | 30 / 30 (100%) | 11 / 11 (100%) | |
| **Depression** | 30 / 30 (100%) | 10 / 11 (91%) | 0.3 |
| **Autism** | | | |
| Yes | 30 / 30 (100%) | 11 / 11 (100%) | |
| **Other liver disease** | | | |
| Yes | 30 / 30 (100%) | 11 / 11 (100%) | |
| **GI surgery** | | | |
| Yes | 30 / 30 (100%) | 11 / 11 (100%) | |
| **Abdominal surgery** | 26 / 30 (87%) | 7 / 11 (64%) | 0.2 |
| **HIV** | | | |
| Yes | 30 / 30 (100%) | 11 / 11 (100%) | |
| **Rheumatic disease** | | | |
| Yes | 30 / 30 (100%) | 11 / 11 (100%) | |
| **No medication** | 18 / 30 (60%) | 7 / 11 (64%) | >0.9 |
| **Metformin** | | | |
| Yes | 30 / 30 (100%) | 11 / 11 (100%) | |
| **Other diabetes medication** | | | |
| Yes | 30 / 30 (100%) | 11 / 11 (100%) | |
| **Insulin** | | | |

| **Post-discharge diabetes mellitus** | **No DM** N = 30[1] | **DM** N = 11[1] | **p-value** |
|---|---|---|---|
| Yes | 30 / 30 (100%) | 11 / 11 (100%) | |
| **Proton-pump inhibitors** | 17 / 30 (57%) | 7 / 11 (64%) | 0.7 |
| **Immunosuppressants** | 30 / 30 (100%) | 10 / 11 (91%) | 0.3 |
| **Antidepressants** | 29 / 30 (97%) | 10 / 11 (91%) | 0.5 |
| **Neuroleptics** | | | |
| Yes | 30 / 30 (100%) | 11 / 11 (100%) | |
| **Paracetamol** | 27 / 30 (90%) | 11 / 11 (100%) | 0.6 |
| **NSAID** | 24 / 30 (80%) | 9 / 11 (82%) | >0.9 |
| **Opiates** | 26 / 30 (87%) | 11 / 11 (100%) | 0.6 |
| **Laxatives** | 29 / 30 (97%) | 11 / 11 (100%) | >0.9 |
| **Statins*** | 23 / 30 (77%) | 8 / 11 (73%) | >0.9 |
| **Ethnicity** | | | >0.9 |
| Asian | 1 / 30 (3.3%) | 0 / 11 (0%) | |
| Caucasian | 29 / 30 (97%) | 11 / 11 (100%) | |
| **Alcohol*** | | | 0.9 |
| Current chronic alcohol consumption | 6 / 30 (20%) | 2 / 11 (18%) | |
| Former chronic alcohol consumption | 3 / 30 (10%) | 2 / 11 (18%) | |
| No alcohol consumption | 21 / 30 (70%) | 7 / 11 (64%) | |
| **Gender** | | | >0.9 |
| Female | 13 / 30 (43%) | 5 / 11 (45%) | |
| Male | 17 / 30 (57%) | 6 / 11 (55%) | |
| **Diet** | | | 0.6 |
| Omnivorous | 27 / 30 (90%) | 11 / 11 (100%) | |
| Vegetarian | 3 / 30 (10%) | 0 / 11 (0%) | |
| **Antibiotics** | | | 0.7 |
| >6month | 25 / 30 (83%) | 9 / 11 (82%) | |
| 6month | 2 / 30 (6.7%) | 0 / 11 (0%) | |
| current | 2 / 30 (6.7%) | 2 / 11 (18%) | |
| week | 1 / 30 (3.3%) | 0 / 11 (0%) | |
| **Nicotine** | | | >0.9 |
| >10 cig./d | 5 / 30 (17%) | 2 / 11 (18%) | |
| 1-10 cig./d | 1 / 30 (3.3%) | 0 / 11 (0%) | |
| No Smoker | 24 / 30 (80%) | 9 / 11 (82%) | |
| **Etiology*** | | | >0.9 |
| Alcoholic | 6 / 30 (20%) | 3 / 11 (27%) | |
| Biliary | 15 / 30 (50%) | 5 / 11 (45%) | |
| Iatrogenic | 3 / 30 (10%) | 1 / 11 (9.1%) | |
| Idiopathic | 6 / 30 (20%) | 2 / 11 (18%) | |
| **Cholestasis** | 8 / 30 (27%) | 4 / 11 (36%) | 0.7 |
| **Age*** | 60 (19) | 59 (22) | 0.7 |
| **BMI** | 28.8 (5.4) | 26.4 (3.3) | 0.3 |
| **Admission Creatinine (mg/dl)** | 1.06 (0.66) | 0.89 (0.38) | 0.4 |
| Unknown | 0 | 1 | |

[1]n / N (%); Mean (SD)

**Suppl. Table 46:** Confounder distribution in matched rectal samples from patients with and without the development of a post-discharge diabetes mellitus (DM). Differences were calculated using Fisher´s exact test, Chi-squared test or Mann-Whitney-U test. BMI – body mass index, GI – gastrointestinal, HIV – human immunodeficiency virus, MASLD – metabolic dysfunction-associated steatotic liver disease, NSAID – nonsteroidal anti-inflammatory drug, SD – standard deviation. *marked variables were used for matching.

| **Species** | **Cohort** | **Method** | **p value MaAsLin2** | **p value LEfSe** |
|---|---|---|---|---|
| *Prevotella nigrescens* | No post-discharge DM | MaAsLin2 & LEfSe | 0.025 | 0.03 |
| *Blautia luti* | No post-discharge DM | MaAsLin2 & LEfSe | 0.025 | 0.029 |
| *Blautia obeum* | No post-discharge DM | MaAsLin2 & LEfSe | 0.031 | 0.045 |
| *Hoylesella buccalis* | No post-discharge DM | MaAsLin2 & LEfSe | 0.01 | 0.006 |
| *Fenollaria massiliensis* | No post-discharge DM | MaAsLin2 & LEfSe | 0.003 | 0.002 |
| *Mobiluncus massiliensis* | No post-discharge DM | MaAsLin2 & LEfSe | 0.017 | 0.013 |
| *Anaerococcus vaginalis* | No post-discharge DM | LefSe | | 0.047 |
| *Varibaculum prostatecancerukia* | No post-discharge DM | LefSe | | 0.045 |
| *Enterococcus faecalis* | Post-discharge DM | MaAsLin2 & LEfSe | <0.001 | 0.009 |
| *Thomasclavelia ramosa* | Post-discharge DM | MaAsLin2 | 0.038 | |
| *Bacteroides nordii* | Post-discharge DM | MaAsLin2 | 0.019 | |

**Suppl. Table 47:** Differential abundance between patients with and without post-discharge diabetes mellitus (DM) was calculated with Linear discriminant analysis effect size (LEfSe) and Microbiome Multivariable Association with Linear Models (MaAsLin2). Median abundance was >0.05%.
